# Evaluation of ATN_PD_ framework and biofluid markers to predict cognitive decline in early Parkinson’s disease

**DOI:** 10.1101/2023.04.21.23288930

**Authors:** Katheryn A.Q. Cousins, David J. Irwin, Thomas F. Tropea, Emma Rhodes, Jeffrey S. Phillips, Alice S. Chen-Plotkin, Michael C. Brumm, Christopher S. Coffey, Ju Hee Kang, Tanya Simuni, Tatiana Foroud, Arthur W. Toga, Caroline M. Tanner, Karl Kieburtz, Brit Mollenhauer, Douglas R. Galasko, Samantha Hutten, Daniel Weintraub, Andrew Siderowf, Kenneth Marek, Gwendlyn Kollmorgen, Kathleen L. Poston, Leslie M. Shaw, The Parkinson’s Progression Marker Initiative

**Affiliations:** Department of Neurology, Perelman School of Medicine, University of Pennsylvania; Department of Biostatistics, College of Public Health, University of Iowa, Iowa City, IA, USA; Department of Pharmacology & Clinical Pharmacology, Inha University, Incheon, South Korea; Feinberg School of Medicine, Northwestern University, Chicago, IL, USA; Department of Medical and Molecular Genetics, Indiana University, Indianapolis, IN, USA; Laboratory of Neuro Imaging, University of Southern California, Los Angeles, CA, USA; Department of Neurology, Weill Institute for Neurosciences, University of California San Francisco, San Francisco, CA, USA; Department of Neurology, University of Rochester Medical Center, Rochester, NY, USA; Department of Neurology, University Medical Center, Göttingen, Paracelsus-Elena-Klinik, Kassel and Scientific Employee with an Honorary Contract at German Center for Neurodegenerative Diseases (DZNE), Göttingen, Germany; Department of Neurology, University of California San Diego, San Diego, CA, USA; The Michael J. Fox Foundation, New York, NY, USA; Department of Psychiatry Perelman, School of Medicine at the University of Pennsylvania, Philadelphia, PA, USA; Michael J. Crescenz VA Medical Center, Parkinson’s Disease Research, Education, and Clinical Center, Philadelphia, PA, USA; Institute for Neurodegenerative Disorders, New Haven, CT, USA; Roche Diagnostics GmbH, Penzberg, Germany; Department of Neurology, Stanford University, Palo Alto, CA, USA; Department of Pathology and Laboratory Medicine, Perelman School of Medicine, University of Pennsylvania

## Abstract

**Background and Objectives:** In Parkinson’s disease (PD), Alzheimer’s disease (AD) co-pathology is common and clinically relevant. However, the longitudinal progression of AD cerebrospinal fluid (CSF) biomarkers – β-amyloid 1-42 (Aβ_42_), phosphorylated tau 181 (p-tau_181_) and total tau (t-tau) – in PD is poorly understood, and may be distinct from clinical AD. Moreover, it is unclear if CSF p-tau_181_ and serum neurofilament light (NfL) have added prognostic utility in PD, when combined with CSF Aβ_42_. First, we describe longitudinal trajectories of biofluid markers in PD. Second, we modified the AD β-amyloid/tau/neurodegeneration (ATN) framework for application in PD (ATN_PD_) using CSF Aβ_42_ (A), p-tau_181_ (T), and serum NfL (N), and tested ATN_PD_ prediction of longitudinal cognitive decline in PD.

**Methods:** Participants were selected from the Parkinson’s Progression Markers Initiative (PPMI) cohort, clinically-diagnosed with sporadic PD or as normal Controls, and followed annually for 5 years. Linear mixed effects models (LMEM) tested the interaction of diagnosis with longitudinal trajectories of analytes (log-transformed, FDR-corrected). In PD, LMEMs tested how baseline ATN_PD_ status (AD [A+T+N±] *vs*. not) predicted clinical outcomes, including Montreal Cognitive Assessment (MoCA; rank-transformed, FDR-corrected).

**Results:** Participants were 364 PD and 168 Controls, with comparable baseline mean (±SD) age (PD=62±10; Control=61±11]; Mann-Whitney-Wilcoxon: *p*=0.40) and gender distribution (PD=231 males [63%]; Control=107 males [64%]; chi-square: *p*=1.0). PD had overall lower CSF p-tau_181_ (β=-0.16, 95%CI=-0.23 – -0.092, *p*=2.2e-05) and t-tau than Controls (β=-0.13, 95%CI=-0.19 – -0.065, *p*=4.0e-04), but not Aβ_42_ (*p*=0.061) or NfL (*p*=0.32). Over time, PD had greater increases in serum NfL than Controls (β=0.035, 95%CI=0.022 – 0.048, *p*=9.8e-07); PD slopes did not differ from controls for CSF Aβ_42_ (*p*=0.18), p-tau_181_ (*p*=1.0) or t-tau (*p*=0.96). Using ATN_PD_, PD classified as A+T+N± (n=32; 9%) had consistently worse cognitive decline, including on global MoCA (β=-73, 95%CI=-110 – -37, *p*=0.00077), than all other ATN_PD_ statuses including A+ alone (A+T-N-; n=75; 21%).

**Discussion:** In early PD, CSF p-tau_181_ and t-tau were low compared to Controls and did not increase over 5 year follow-up. Even so, classification using modified ATN_PD_ (incorporating CSF p-tau_181_ with CSF Aβ_42_ and serum NfL) may identify biologically-relevant subgroups of PD to improve prediction of cognitive decline in early PD.

## 1. Introduction

Parkinson’s disease (PD) is a progressive movement disorder associated with underlying α-synuclein positive Lewy bodies^1^. Clinically-relevant levels of Alzheimer’s disease (AD) pathology are common in Lewy body disease (LBD), with roughly 40% having sufficient β-amyloid plaques and tau neurofibrillary tangles for a secondary neuropathological diagnosis of intermediate-to-high AD neuropathologic change (ADNC)^2, 3^. Moreover, AD co-pathology in PD may contribute to more severe episodic memory and language impairments, as well as worse postural instability and shorter survival^3–6^. AD biomarkers may prove important to prognosticate clinical outcomes in PD. It is therefore important to track when AD pathology might arise in PD, yet there have been limited studies of AD biomarker trajectories in early PD.

In addition to poorly understood biomarker dynamics in PD, there is no established framework for AD biomarker classification in PD. The ATN framework was developed to diagnose individuals along a continuum of AD^7^, and cerebrospinal fluid (CSF) biomarkers can determine likely presence (+) or absence (-) of AD pathological hallmarks: brain accumulation of β-amyloid (A), tau (T), and/or neurodegeneration (N). Typically, low CSF β-amyloid 1-42 (Aβ_42_) indicates A+, and conversely high phosphorylated tau 181 (p-tau_181_) and total tau (t-tau) in CSF can indicate T+ and N+, respectively. More recently, neurofilament light chain (NfL) has been used as a marker of N status, instead of t-tau. Indeed, accumulating evidence shows that serum NfL may be a useful marker of disease progression and degeneration in PD^8–11^. While the the ATN framework is starting to be applied in LBD^12, 13^, its prognostic utility has not been thoroughly evaluated in PD. In PD, NfL is untested in the context of ATN, and it is unknown if there will be added prognostic value when combined with AD CSF biomarkers. There is also evidence that CSF p-tau_181_ and t-tau are less informative in PD, showing an altered profile and low levels^14^ that may reduce their sensitivity for classifying AD^15^. Thus, CSF Aβ_42_ alone may be sufficient to detect evidence of AD pathology in PD^16^ and to predict clinical decline in PD^17, 18^. The added utility of CSF p-tau, as well as serum NfL, when combined with CSF Aβ_42_ (*i.e.*, ATN) is undetermined, and head-to-head comparisons of AD biomarker strategies to predict clinical cognitive decline in PD are lacking.

This study of the Parkinson’s Progression Marker Initiative (PPMI) cohort had two goals: to track the potentially unique AD biofluid trajectories in PD, and to test how biomarker strategies may be applied to identify biologically-relevant subgroups and improve clinical prognosis in PD. In part 1, we described longitudinal CSF (Aβ_42_, p-tau_181_, t-tau) and serum NfL biofluid trajectories from baseline to year 5 in PD, compared to clinically-normal Controls without motor or cognitive impairments. We also tested if biomarker trajectories in PD differed by baseline CSF Aβ_42_ (*i.e.*, evidence of β-amyloid accumulation) or baseline CSF α-synuclein (α-syn) (*i.e.*, potential evidence of α-syn accumulation). In part 2, we compared two biomarker strategies to predict future clinical outcomes in PD: 1) CSF Aβ_42_ as a marker of A status and 2) a modified ATN strategy (ATN_PD_) using a lower PD-established cutpoint for T status^19^ and serum NfL as the marker of N. Ancillary analyses evaluated traditional ATN (CSF Aβ_42_, p-tau_181_, and t-tau) to test if results were cutpoint or analyte dependent. We tested the hypothesis that ATN_PD_ incorporating T and N status would improve prediction of clinical outcomes in PD; alternatively A status alone may be sufficient for prognosis in early PD. We also predicted that AD biomarker strategies would specifically associate with declines in cognition, language, and postural stability in PD, but not with other motor and autonomic functioning, in line with previous literature^3–6^.

## 2. Materials and methods

PPMI is a longitudinal, observational study, with data collection from approximately 50 international sites. Data were selected on September 7, 2021 from the PPMI database^20, 21^. All procedures were performed with prior approval from ethical standards committees at each participating institution and with informed consent from all study participants. CSF and clinical data were obtained annually at baseline (year 0) and follow-up visits (years 1 – 5); serum NfL was obtained at years 0, 1, 2, 3, and 5. A subset of these data was previously reported^17^.

### 2.1 Participants

Inclusion criteria were participants diagnosed with PD who, at baseline, were drug-naïve and had CSF Aβ_42_, p-tau_181_, t-tau, α-syn, and serum NfL measurements (n=364); in addition they were within 2 years of PD diagnosis, not diagnosed with dementia, and sporadic (without *LRRK2*, *GBA*, or *SNCA* mutations). Controls were included as a reference group who had a normal clinical diagnosis (n=168). Exclusion criteria were prodromal PD (conversion to PD during the follow-up period), parkinsonism but without evidence of dopaminergic deficit syndrome, or a diagnosis change to multiple system atrophy (MSA) or corticobasal syndrome. Of those followed at baseline, 284 PD (78%) and 141 Controls (84%) were still followed by year 5.

### 2.2 CSF and serum assays

Biofluid collection and handling were completed according to standard operating procedures at each institution (see biologics manual: <ppmi.info.org>). CSF α-syn and serum NfL were analyzed as previously described^8, 22^. For measurement of CSF Aβ_42_, p-tau_181_, and t-tau, CSF samples were shipped from the PPMI Biorepository Core Laboratories to the Penn Biomarker Research Laboratory using Elecsys® electrochemiluminescence immunoassays on the cobas e 601 analysis platform (Roche Diagnostics)^23, 24^.

Biofluid data were from two batches (2016, 2021). The 2021 batch was 68 analytical runs done in a total of 34 days: 2/22/2021 through 4/16/2021. Single reagent lots were used throughout the study for each analyte.

A total of 2733 CSF samples were processed in this project, including 2463 PPMI patient samples, 134 PPMI pools, and 136 internal UPenn pool controls. Among the 2463 patient samples, there were 75 “bridging” samples previously analyzed in the 2017 batch. These 75 retests were planned for the purpose of evaluating the assay lot-to-lot performance.

The “bridging” samples were mixed-in with the regular study samples during the first 10 days of analyses (first 20 runs). Regression equations were obtained using the Passing-Bablok method^25^, using the regression equation: *[2021 result] =* β*_0_ +* β*_1_[2017 result]*.

Summarized are the model intercepts (β_0_) and 95% confidence intervals (CI) for each analyte: Aβ_42_=8.04 (95%CI=-169.9 – 153.4); t-tau=-3.29 (95%CI=-14.76 – 8.23); p-tau_181_=0.36 (-0.18 – 1.01)]. The summarized model slopes (β_1_) are Aβ_42_=1.15 (95%CI=0.99 – 1.40); t-tau=1.10 (95%CI=0.94 – 1.10); p-tau_181_=0.93 (95%CI=0.88 –0.97). Intercepts and slopes for t-tau and p-tau_181_ met criteria for acceptable lot-to-lot performance: intercept <50% lower than the lowest reportable value, and slope between 0.90–1.10. To achieve complete comparability of the Aβ_42_ assay, we re-scaled the results of all CSF samples to the values measured in 2017, using the regression equation for Aβ_42_.

Precision performance was evaluated using 4 CSF pools, each included in the 68 analytical runs. For Aβ_42_ %CV values ranged from 3.8 to 5.8%; t-tau %CV from 2.9 to 3.3% and for p-tau_181_ %CV from 2.8 to 5.2%.

#### 2.2.1 ATN framework and interpretation

##### A status

CSF Aβ_42_ determined A status for all strategies. To account for CSF Aβ_42_ pre-analytical factors that affect cutoff application in the PPMI dataset^23, 26^, a threshold of 683 was calculated^19^: CSF Aβ_42_≤683 pg/mL indicated A+ and Aβ_42_>683 pg/mL indicated A-for all strategies (A Status, modified ATN_PD_, and traditional ATN).

##### Modified ATN_PD_

As above, CSF Aβ_42_≤683 pg/mL indicated A+. For T status, CSF p-tau_181_≥13 pg/mL indicated T+ based on a PD-established cutpoint^19^. Serum NfL determined N status. Because no established thresholds were available for serum NfL, the threshold was determined from this PD sample based on the 75% quartile for NfL at baseline; thus NfL≥19.05 pg/mL indicated N+.

##### Traditional ATN

Traditional ATN applied thresholds established in AD. As above, CSF Aβ_42_≤683 pg/mL indicated A+. For T status, CSF p-tau_181_≥24 pg/mL indicated T+^27^. For N status, CSF t-tau≥266 indicated N+^27^.

##### ATN_PD_ and ATN Interpretation

Despite different biomarkers and thresholds, both ATN_PD_ and ATN used the same schema to interpret statuses^7^. Combining biomarkers classified individuals along a spectrum of AD: normal (A-T-N-), early Alzheimer’s change (A+T-N-), AD (A+T+N-, A+T+N+), suspected non-AD pathology (SNAP) (A-T+N-, A-T-N+, A-T+N+), and concomitant Alzheimer’s change and SNAP (A+T-N+). In sum, both A+ and T+ are required to meet a diagnosis of AD, while A-in the presence of T+ and/or N+ is considered SNAP.

### 2.3 Demographics, Clinical, and Genetic Data

Demographic variables were age at baseline visit, age at symptom onset, disease duration (years from onset to test), education (years), and gender. The PPMI genetics core analyzed blood samples for apolipoprotein E (APOE) genotype^28^ and mutations associated with PD risk (*e.g.*, *LRRK2*, *GBA*, *SNCA*)^29^.

Clinical variables were obtained for each visit (baseline and follow-ups). Metrics of cognitive function were Montreal Cognitive Assessment (MoCA; lower scores indicate worse global function), Hopkins Verbal Learning Test (HVLT) recognition discrimination score (lower scores indicate worse episodic memory), Symbol Digit Modalities Test (SDMT; lower scores indicate worse processing speed), Benton Judgment of Line Orientation score (JOLO; lower scores indicate worse visuospatial functioning), Semantic Fluency (lower scores indicate worse verbal generativity and semantic knowledge), Letter Number Sequencing (lower scores indicate worse executive functioning), and the Movement Disorders Society modified Unified Parkinson’s disease rating scale (MDS-UPDRS) part I item 1.1 Cognitive Impairment (MDS-UPDRS I Cog; higher scores indicate more severe cognitive impairment)^30^. Metrics of motor functioning were Hoehn & Yahr^31^, MDS-UPDRS part III total motor score off levodopa treatment (MDS-UPDRS III Motor)^30^, tremor, and postural instability/gait dysfunction (PIGD) (higher scores indicate more severe impairment). Tremor and PIGD were calculated averages derived from MDS-UPDRS part II and III relevant items^32^. The metric for non-motor/cognitive autonomic functioning was the Scales for Outcomes in Parkinson’s Disease-Autonomic Dysfunction (SCOPA-AUT; higher scores indicate more severe impairment)^33^.

### 2.4 Statistical Analyses

Biofluid, demographic, and clinical variables had a non-normal distribution. Mann– Whitney–Wilcoxon tests performed pairwise comparisons for continuous variables, and chi-square tests compared frequency distributions of categorical variables. In models, CSF and serum biomarkers were log-transformed, and β estimates and 95% confidence intervals (95% CI) are reported. Because distributions varied for clinical variables, clinical variables were rank-transformed to perform non-parametric analyses^34^. Missing data were dropped listwise from models. Statistical tests were performed with a significance threshold of α=0.05. Nominal and false detection rate (FDR)-adjusted *p*-values are reported. Analyses were conducted using R version 4.1.2 (2021-11-01) statistical software.

#### 2.4.1 Part 1: Biofluid Trajectories in PD and Controls

Linear mixed effects models (LMEMs) tested how each analyte changed over time (Equation 1); an interaction term between visit year (0-5) and group tested for divergence in analyte trajectories between PD and Controls. Models covaried for age at baseline, gender, APOE ε4 (dominant model: 0 *vs*. 1/2 ε4 alleles), with a random intercept for individual (γ) and residual error (ε).

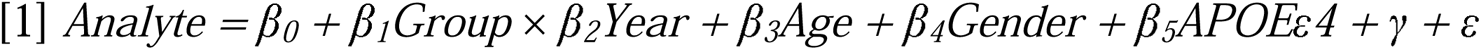

In PD, we tested how evidence of β-amyloid at baseline related to longitudinal trajectories in p-tau_181_, t-tau, and NfL; LMEMs tested for an interaction between time and baseline CSF Aβ_42_ (Equation 2). Models covaried for age at baseline, gender, and APOE ε4 carrier status.

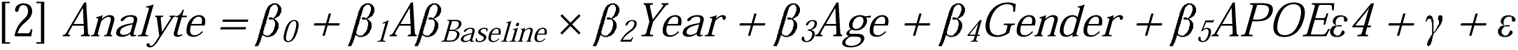

Longitudinal CSF α-syn trajectories in this PPMI cohort were previously reported^22^. In addition to baseline CSF Aβ_42_, we also tested how baseline CSF α-syn interacted with longitudinal trajectories in CSF Aβ_42_, p-tau_181_, t-tau, and serum NfL (Equation 3).

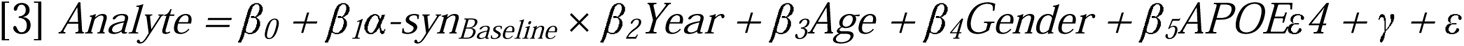

We examined distributions of 2 biomarker strategies to classify PD over time: modified ATN_PD_ and A status. Chi-square tests examined if classifications changed from year 0 to year 5. We included traditional ATN as a reference.

#### 2.4.3 Part 2: Prediction of clinical outcomes by biomarker strategy

We compared ATN_PD_ to A status to predict longitudinal clinical outcomes. We tested if declines in clinical outcomes were associated with co-occurring AD in PD, as defined by ATN_PD_ and by A status. LMEMs tested for an interaction between ATN_AD_ (AD *vs*. other statuses) and year (Equation 4); we also tested for an interaction between A status (A+ *vs*. A-) and year (Equation 5). Models covaried for education and gender, as these variables are known to influence cognitive performance.

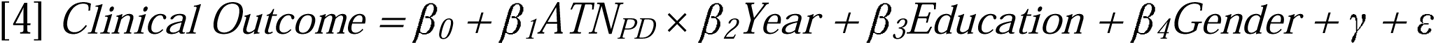

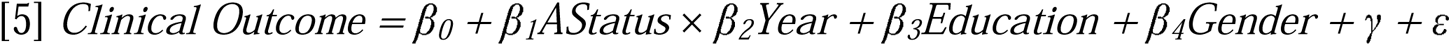

To determine added utility of CSF p-tau_181_ and serum NfL when combined with CSF Aβ_42_, analyses of variance (ANOVA) compared model fitness between nested models: A status interacting with time (Equation 5) compared to AT (Equation 6), and AT compared to ATN (Equation 7). Metrics of fitness were Akaike information criterion (AIC) and log likelihood; chi-square and *p*-values indicated if the model with more parameters significantly reduced variance.

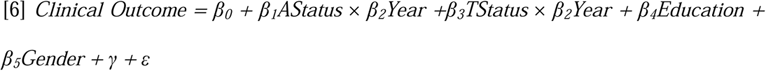

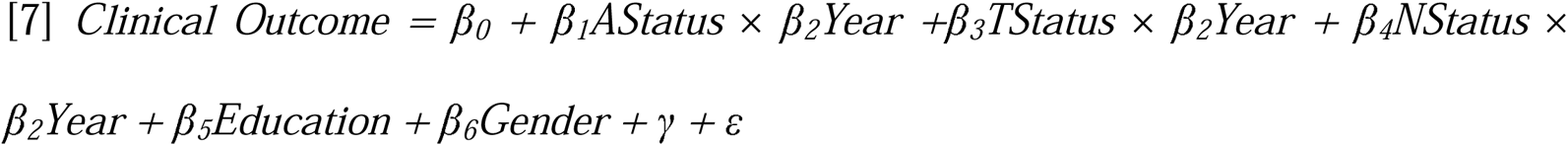

### 2.5 Data Availability

Data contained in this study were obtained from the PPMI database and are available upon request at www.ppmi-info.org.

### 2.6 Standard Protocol Approvals, Registrations, and Patient Consents

Informed consent was obtained from all participants or legal representatives under guidelines established and approved by the ethical standards committees at each participating institution.

## 3. Results

Table 1 summarizes characteristics of PD and Control cohorts at baseline. Supplemental Figure 1 illustrates pairwise correlations of analytes at baseline across PD and Controls; CSF Aβ_42_, p-tau_181_, t-tau, and α-syn were positively correlated with each other in both groups (all *p*<0.0001). Serum NfL was not correlated with either CSF Aβ_42_ or CSF α-syn at baseline (all *p*>0.31).

**Table 1:**
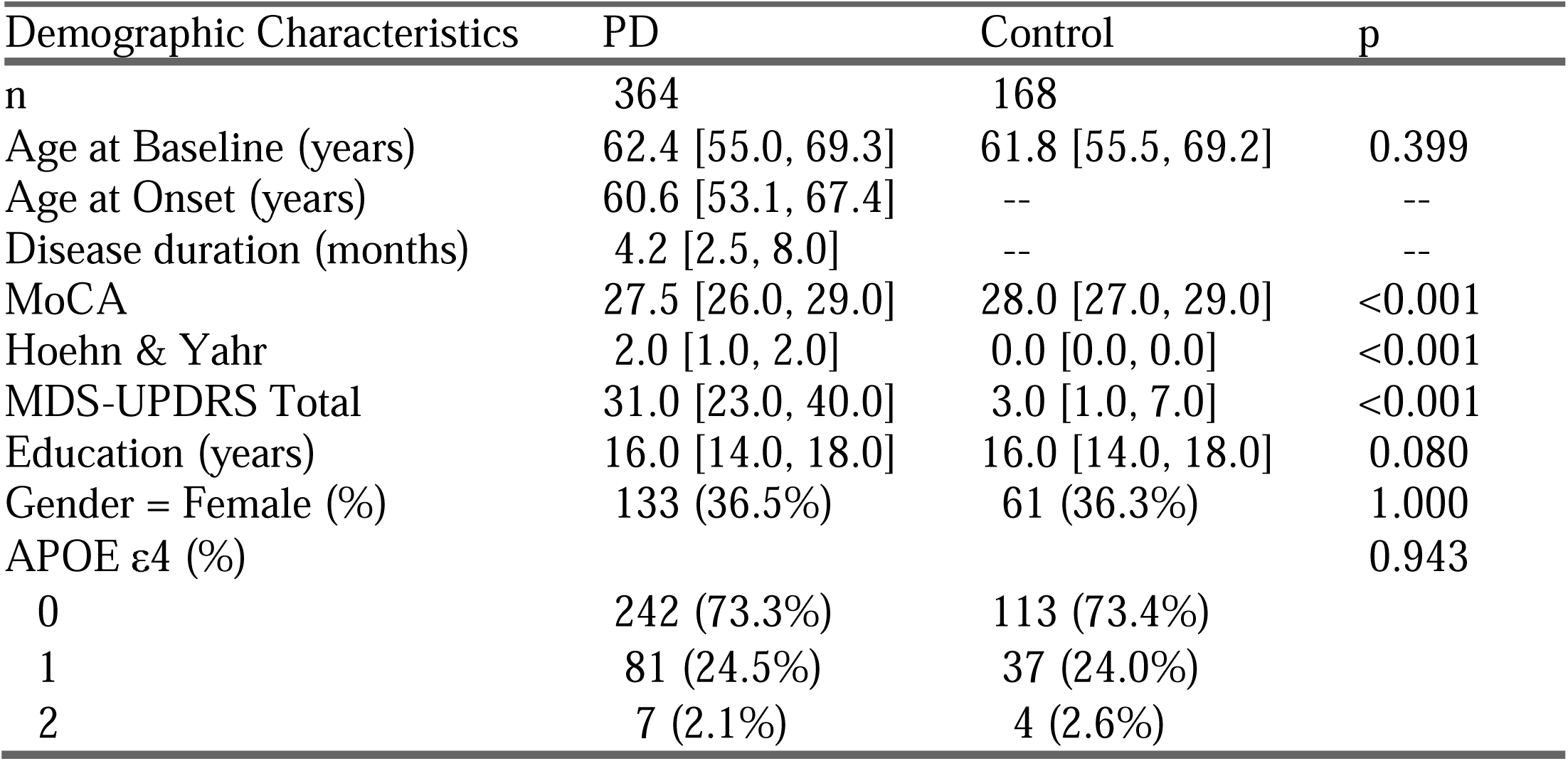
Demographic, genetic, and clinical characteristics of PD and Controls. For continuous variables, median and interquartile range (IQR) are reported; Wilcoxon tests performed group comparisons. For categorical variables, count (percentage [%]) are provided; chi-square tests performed frequency comparisons. p-values are reported for group comparisons.

### Missing values at baseline

APOE ε4 was missing for 34 PD and 14 Controls. PD were missing data for HVLT (n=1), Semantic Fluency (n=1), Letter Number Sequencing (n=1), JOLO (n=1), and SCOPA-AUT (n=8).

### Longitudinal Retainment

By year 5, 284 (78% of baseline) PD had been assessed for MDS-UPDRS I Cog, PIGD, and Tremor; 283 (79% of baseline) had completed SCOPA-AUT; 282 (78% of baseline) PD had completed HVLT, Letter Number Sequencing, MoCA, Semantic Fluency, and SDMT; 280 (77% of baseline) had completed JOLO; 209 (57% of baseline) had completed Hoehn & Yahr; and 207 (57% of baseline) had completed MDS-UPDRS III Motor.

### 3.1 Part 1: Longitudinal biofluid trajectories

First, LMEMs tested longitudinal trajectories of each analyte and for a group by time interaction (Figure 1). PD showed greater increases in serum NfL over time than Controls (β=0.035, 95%CI=0.022 – 0.048, *p*=2.5e-07), and applying FDR-correction (FDR-*p*=9.8e-07). Greater declines in CSF Aβ_42_ in PD compared to Controls (β=-0.01, 95%CI=-0.02 – -2e-04, *p*=0.046) did not survive FDR-correction (FDR-*p*=0.18). There was no significant interaction of group with longitudinal trajectories of p-tau_181_ (*p*=0.30) or t-tau (*p*=0.24).

**Figure 1.**
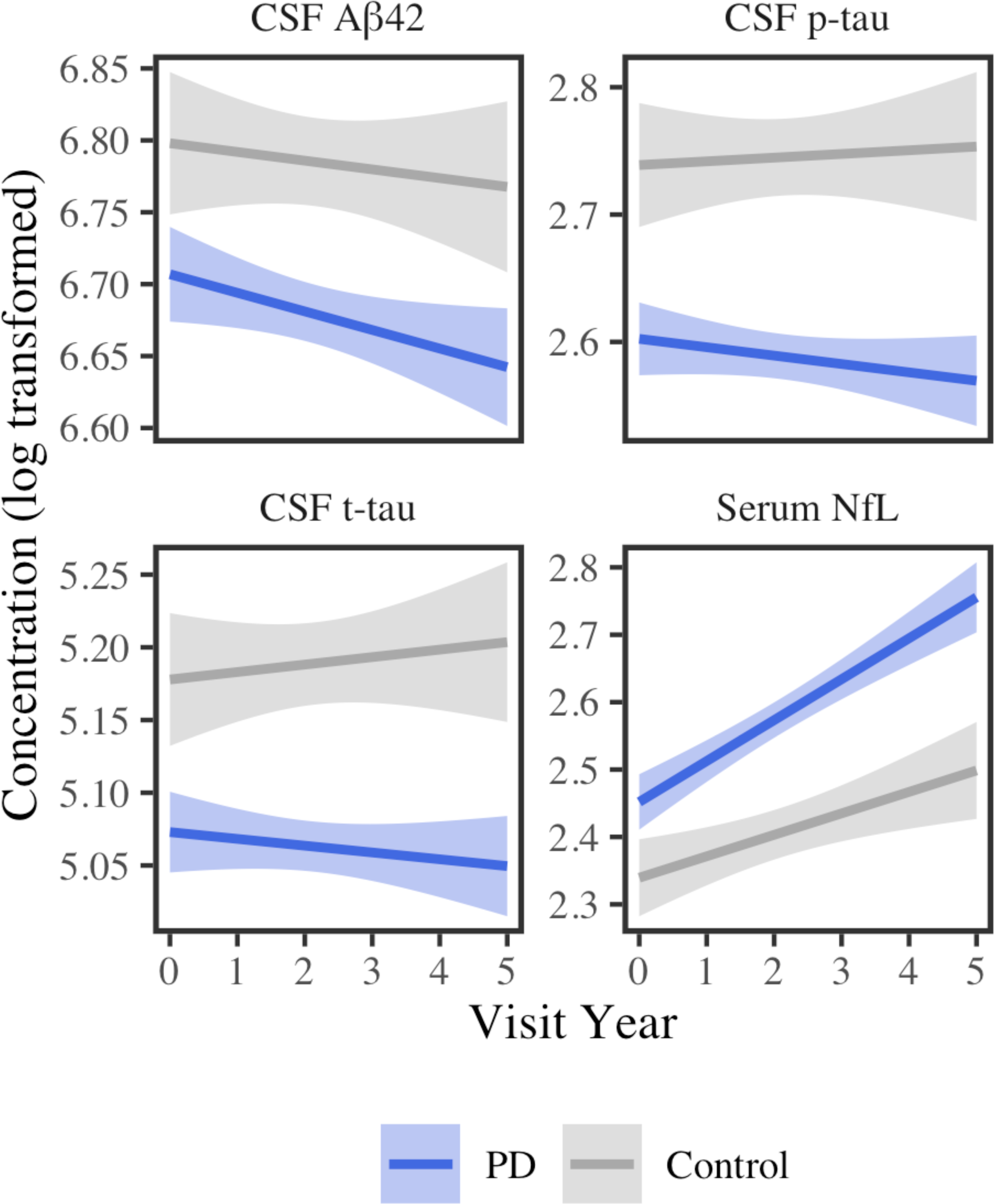
Analyte trajectories over time. Least squares regression lines and standard error are plotted for PD (blue) and Controls (grey) for each analyte by visit year (0-5).

Testing main effect of diagnosis group, PD had overall lower CSF p-tau_181_ (β=-0.16, 95%CI=-0.23 – -0.092, *p*=5.5e-06) and t-tau (β=-0.13, 95%CI=-0.19 – -0.065, *p*=1e-04) relative to controls, and survived FDR-correction (p-tau_181_ FDR-*p*=2.2e-05; t-tau FDR-*p*=4.0e-04). Lower CSF Aβ_42_ in PD (β=-0.091, 95%CI=-0.16 – -0.018, *p*=0.015) did not survive FDR-correction (FDR-*p*=0.061), and group did not show a main effect for serum NfL (β=0.06, 95%CI=-0.0069 – 0.13, *p*=0.080). Full models are reported in Supplementary Table 1.

Within PD, we next tested how longitudinal changes of p-tau_181_, t-tau and NfL related to baseline CSF Aβ_42_ (Figure 2); LMEMs tested for a time by baseline CSF Aβ_42_ (continuous variable) interation. In PD, higher baseline Aβ_42_ was associated with greater declines in p-tau_181_ (β=-0.026, 95%CI=-0.035 – -0.017, *p*=2.5e-08) and t-tau over time (β=-0.025, 95%CI=-0.035 – -0.014, *p*=1.2e-05); both p-tau_181_ (FDR-*p*=7.5e-08) and t-tau (FDR-*p*=3.6e-05) survived FDR-correction. Baseline CSF Aβ_42_ did not affect the trajectory of NfL (*p*=0.73). Full models are reported in Supplementary Table 2.

**Figure 2.**
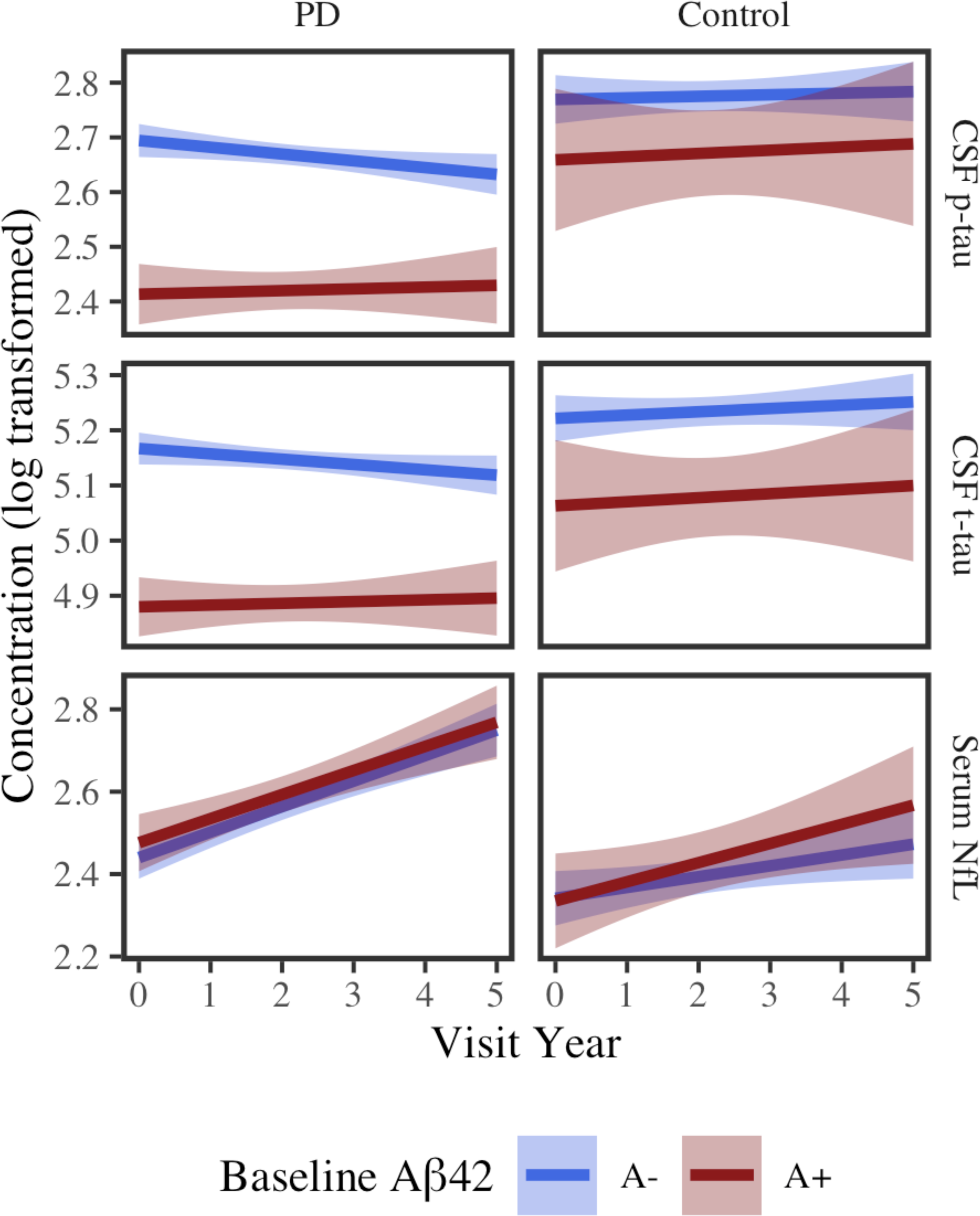
Changes in analytes over time by baseline CSF Aβ_42_. Least squares regression lines with standard error show analyte concentrations over time for PD (left panels) and Controls (right panels). For visualization, baseline CSF Aβ_42_ was dichotomized; color represents low (red; <685) or high (blue; >685) CSF Aβ_42_ at baseline.

Additionally, we tested how longitudinal trajectories of CSF Aβ_42_, p-tau_181_, t-tau and serum NfL related to baseline CSF α-syn. PD biofluid trajectories were not affected by baseline CSF α-syn (all *p*>0.16; data not shown).

#### 3.1.1 PD biomarker classifications over time

Figure 3 shows how each biomarker strategy classified PD patients at baseline and in year 5. For A status, 116 (32%) PD patients were A+ at baseline. Under modified ATN_PD_, 32 (9%) PD patients were classified as AD (A+T+N±) at baseline. Under traditional ATN, 7 (2%) PD patients were classified as AD (A+T+N±) at baseline. Chi-square tests tested if classifications changed over time. Both A status (χ^2^=7, *p*=0.012) and modified ATN_PD_ classifications differed from baseline to year 5 (χ^2^=13, *p*=0.012); traditional ATN classifications did not significantly differ (χ^2^=7.1, *p*=0.067).

**Figure 3.**
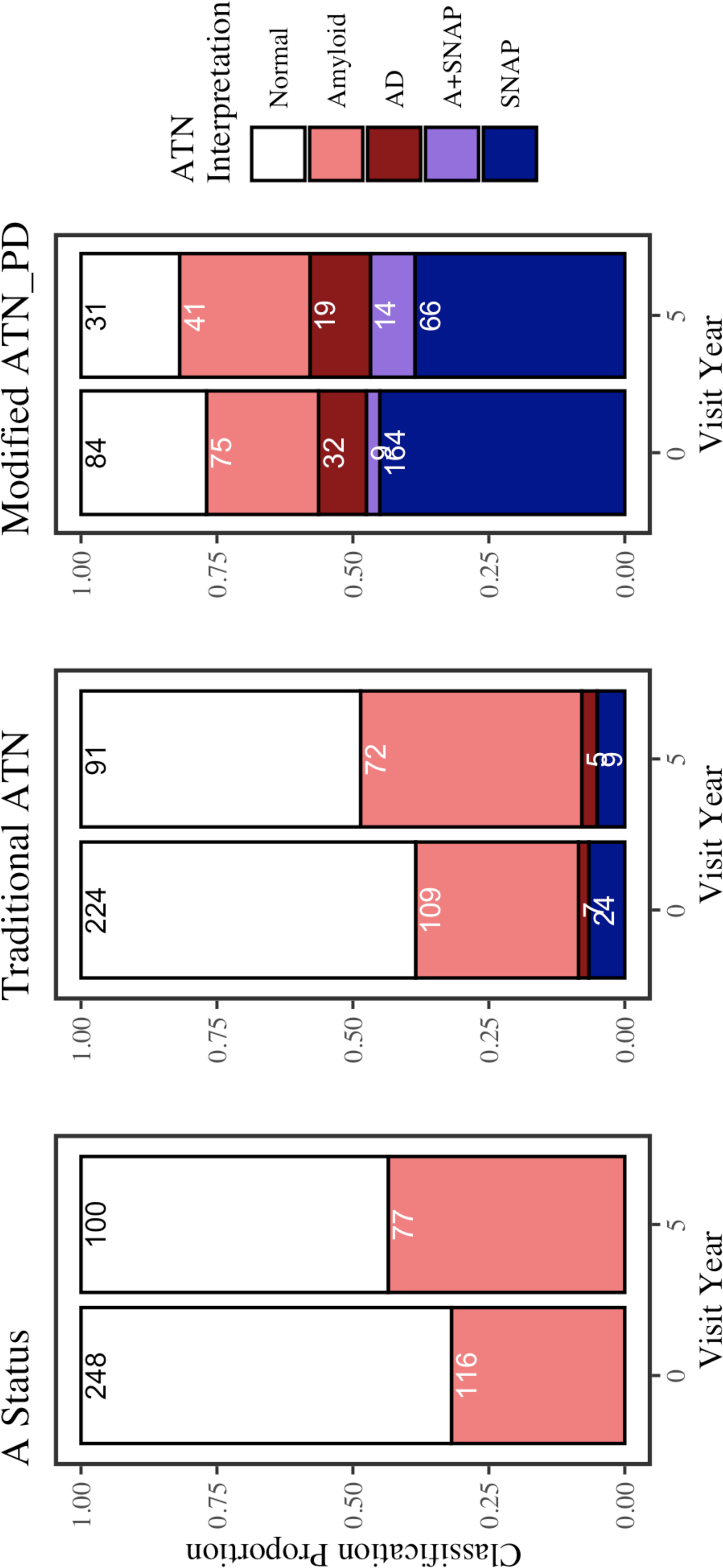
Classifications of PD across biomarker strategies. Classification proportions of PD over time by traditional ATN, A Status, and modified ATN_PD_. Sample sizes for each classification are reported. Color indicates ATN and ATN_PD_ interpretation: Normal (white; A-T-N-), Amyloid (coral; A+T-N-), AD (A+T+N-, A+T+N+), A+SNAP (purple; A+T-N+), and SNAP (blue; A-T+N-, A-T-N+, A-T+N+). For A Status, interpretation is either A-(white) or A+ (coral). Sample sizes for each classification are reported.

### 3.2 Part 2: Biomarker strategies to predict longitudinal clinical outcomes

#### 3.2.1 Longitudinal cognitive outcomes

Next, we tested how modified ATN_PD_ at baseline predicted longitudinal cognitive decline, and how ATN_PD_ performed compared to A status (Figure 4). Full models, as well as traditional ATN results, are reported in the Supplementary Tables 3-5.

**Figure 4.**
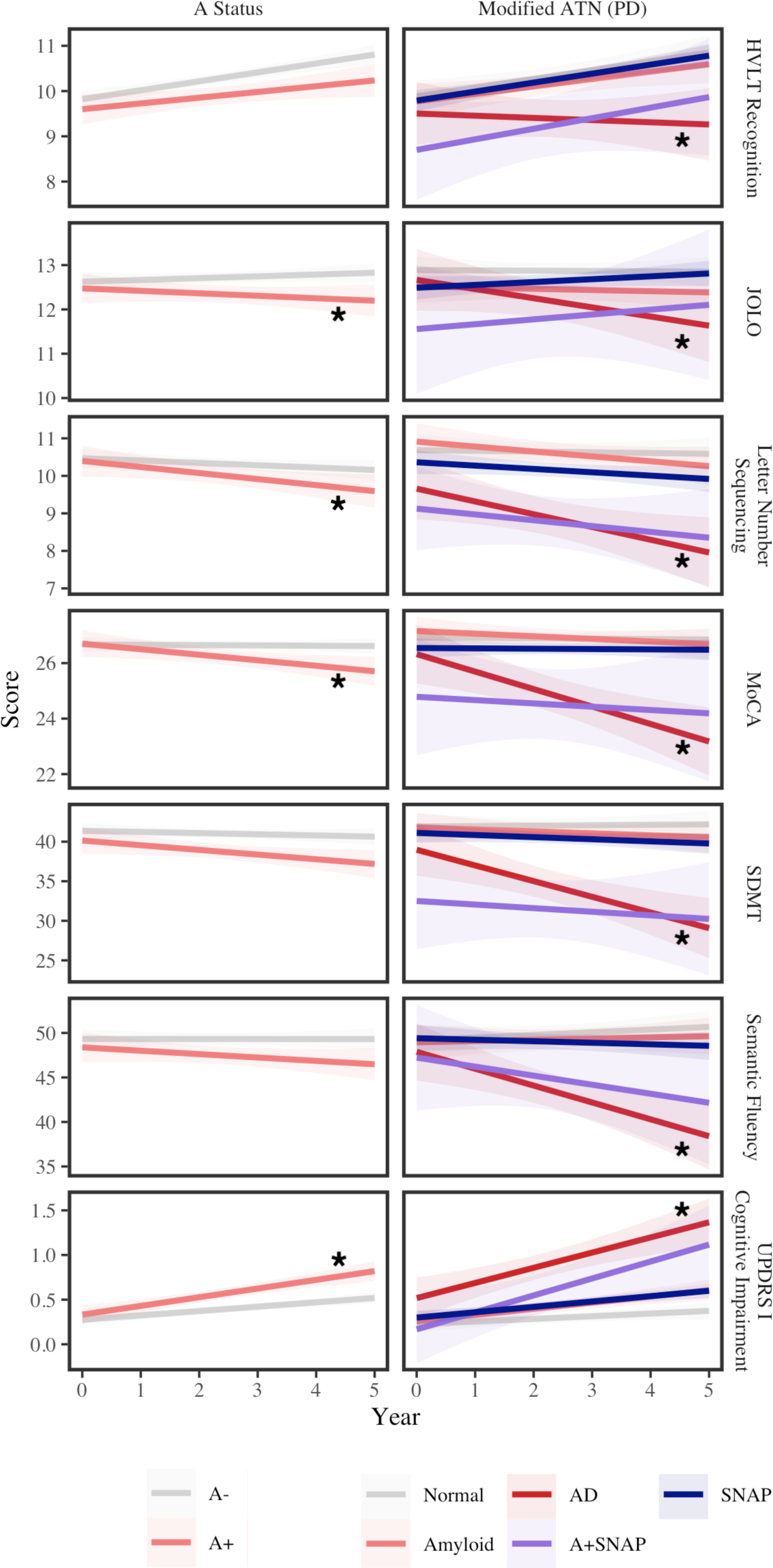
Modified ATN_PD_ *vs*. A status associations with cognitive decline. Least squares regression lines and standard error intervals plot performance over time for each cognitive metric. Color indicates classification by A status (left panels: A-= grey; A+ = coral) and ATN_PD_ (right panels: Normal = grey; Amyloid = coral; AD = red; A+SNAP = purple; SNAP = blue). Asterisks indicate nominal *p*<0.05.

##### ATN_PD_

Testing how AD[A+T+N±] (n=32) *vs*. all other statuses (n=332) at baseline predicts cognitive decline, LMEMs showed a significant interaction between ATN_PD_ and time with all cognitive tests before FDR-correction. A+T+N± PD had more severe declines for MoCA (β=-73, 95%CI=-110 – -37, *p*=6.4e-05; FDR-*p*=0.00077), SDMT (β=-60, 95%CI=-88 – -32, *p*=3.7e-05; FDR-*p*=0.00044), Semantic Fluency (β=-73, 95%CI=-100 – -46, *p*=1.1e-07; FDR-*p*=1.3e-06), and MDS-UPDRS I Cog than all other ATN_PD_ statuses (β=65, 95%CI=30 – 100, *p*=3e-04; FDR-*p*=0.0036). Greater declines for A+T+N± PD in HVLT (β=-58, 95%CI=-100 – -15, *p*=0.0079), JOLO (β=-53, 95%CI=-92 – -13, *p*=0.0089), and Letter Number Sequencing (β=-39, 95%CI=-71 – -7.2, *p*=0.016) did not survive FDR-correction (HVLT FDR-*p*=0.095; JOLO FDR-*p*=0.11; Letter Number Sequencing FDR-*p*=0.20).

##### A Status

We repeated models using A Status to test if A status alone was sufficient to predict cognitive decline. Testing A+ (n=116) *vs*. A-(n=248) at baseline, LMEMs tested for an interaction between A status and time. A+ PD had lower MoCA (β=-31, 95%CI=-51 – -11, *p*=0.0026; FDR-*p*=0.032) and more severe MDS-UPDRS I Cog than A- PD (β=33, 95%CI=13 – 54, *p*=0.0012; FDR-*p*=0.014). Worse JOLO (β=-24, 95%CI=-46 – -1.5, *p*=0.037) and Letter Number Sequencing (β=-19, 95%CI=-37 – -0.22, *p*=0.047) for A+ PD did not survive FDR-correction (JOLO FDR-*p*=0.44; Letter Number Sequencing FDR-*p*=0.57). The interaction was not significant for HVLT (*p*=0.20), SDMT (β=-15, 95%CI=-31 – 1.7, *p*=0.08)0, or Semantic Fluency (β=-15, 95%CI=-31 – 0.63, *p*=0.060).

#### 3.2.2 Longitudinal motor and autonomic outcomes

We tested how modified ATN_PD_ at baseline predicted longitudinal motor/autonomic decline, and how ATN_PD_ performed compared to A status (Figure 5). Full models for motor and autonomic functioning, as well as traditional ATN results, are reported in the Supplementary Tables 6-8.

**Figure 5.**
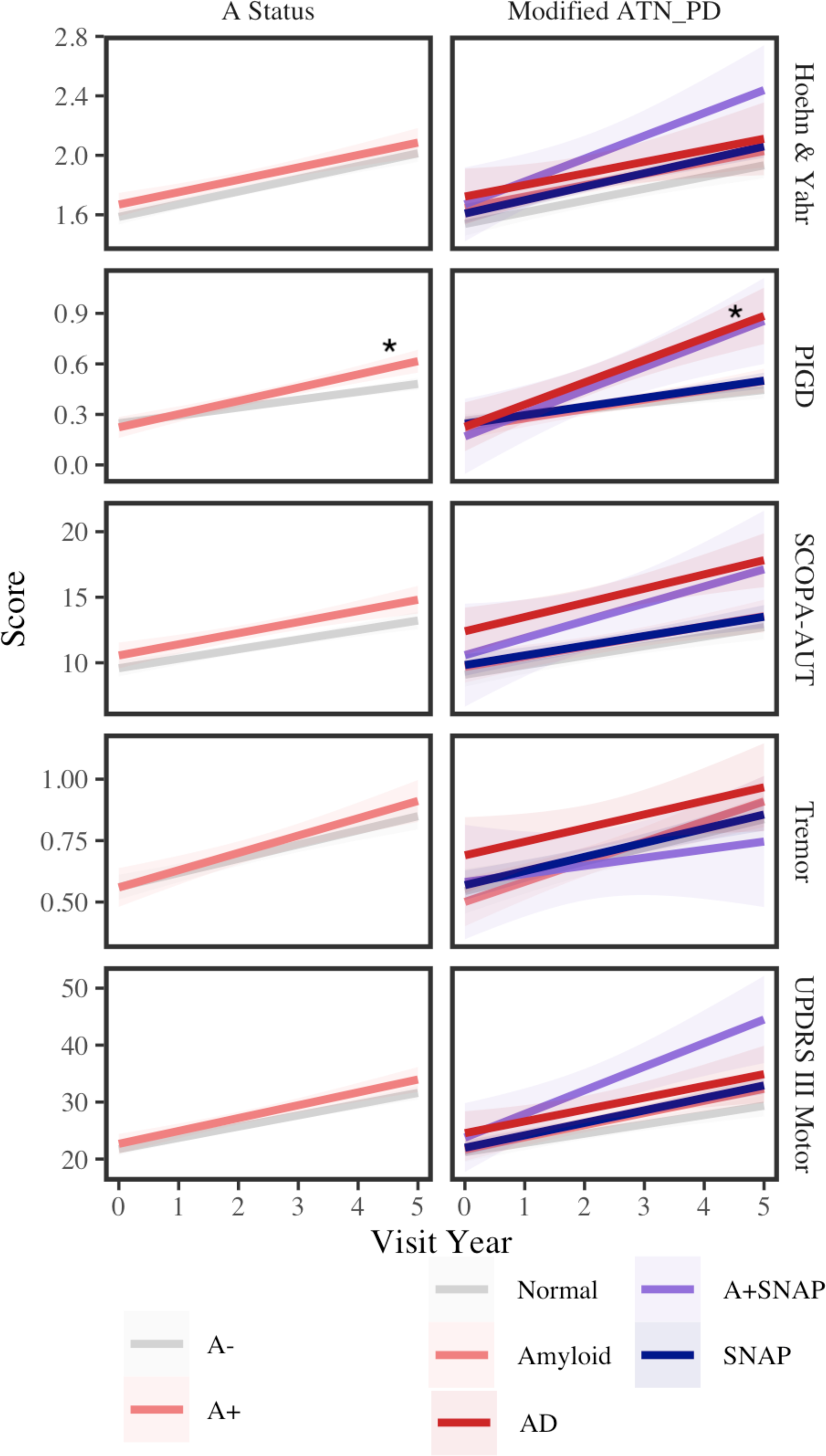
Modified ATN_PD_ *vs*. A status associations with motor and autonomic decline. Least squares regression lines and standard error intervals plot performance over time for each motor/autonomic metric. Color indicates classification by A status (left panels: A- = grey; A+ = coral) and ATN_PD_ (right panels: Normal = grey; Amyloid = coral; AD = red; A+SNAP = purple; SNAP = blue). Asterisks indicate nominal *p*<0.05.

##### ATN_PD_

We tested how AD[A+T+N±] (n=32) *vs*. all other statuses (n=332) at baseline predicts longitudinal motor and autonomic decline. LMEMs showed a significant interaction between ATN_PD_ and time with for PIGD (β=69, 95%CI=29 – 110, *p*=0.00069); this interaction survived FDR correction (FDR-*p*=0.0083). There was not a significant interaction for SCOPA-AUT (β=27, 95%CI=-1.8 – 56, *p*=0.066), UPDRS III Motor (*p*=0.42), Tremor (*p*=0.95), or Hoehn & Yahr (*p*=0.78).

##### A Status

We tested how A+ (n=116) *vs*. A- (n=248) at baseline predicted longitudinal motor and autonomic decline. The interaction between A status and time for PIGD (β=29, 95%CI=6.4 – 52, *p*=0.012) did not survive FDR correction (FDR-*p*=0.15). There was no interaction for SCOPA-AUT (β=15, 95%CI=-1.9 – 32, *p*=0.083), UPDRS III Motor (*p*=0.24), Tremor (*p*=0.11), or Hoehn & Yahr (*p*=0.81).

#### 3.2.3 Model comparison: modified ATN_PD_ vs. A status

To determine if ATN_PD_ improved predictions of clinical decline, ANOVAs compared model fitness of A status *vs*. AT_PD_, and AT_PD_ *vs*. ATN_PD_ (Table 2). Results showed improved model fit for ATN_PD_ for all cognitive outcomes except JOLO, with most metrics seeing additional improvement for both T and N statuses. In addition, there was improved model fit when N status was included for PIGD.

**Table 2:** Comparison of nested models (A vs. AT_PD_ vs. ATN_PD_) for prediction of clinical outcomes. Summary of ANOVAs comparing model fitness for modified ATN_PD_. Columns report number of parameters per model (npar), AIC, log liklihood (logLik), chi-square (χ^2^), degrees of freedom (Df), p-values, and FDR-p.

| | npar | AIC | logLik | $\chi^2$ | Df | p | FDR-p |
| --- | --- | --- | --- | --- | --- | --- | --- |
| HVLT.model = A | 8 | 29,794 | -14,889 |  |  |  |  |
| HVLT.model = AT_PD | 10 | 29,790 | -14,885 | 7.7 | 2 | 0.021 | 0.043 |
| HVLT.model = ATN_PD | 12 | 29,782 | -14,879 | 11.9 | 2 | 0.003 | 0.01 |
| JOLO.model = A | 8 | 29,473 | -14,728 |  |  |  |  |
| JOLO.model = AT_PD | 10 | 29,476 | -14,728 | 0.4 | 2 | 0.819 | 0.819 |
| JOLO.model = ATN_PD | 12 | 29,474 | -14,725 | 6.5 | 2 | 0.038 | 0.066 |
| Letter Number.model = A | 8 | 29,085 | -14,534 |  |  |  |  |
| Letter Number.model = AT_PD | 10 | 29,079 | -14,530 | 9.4 | 2 | 0.009 | 0.024 |
| Letter Number.model = ATN_PD | 12 | 29,074 | -14,525 | 9.0 | 2 | 0.011 | 0.026 |
| MoCA.model = A | 8 | 29,358 | -14,671 |  |  |  |  |
| MoCA.model = AT_PD | 10 | 29,356 | -14,668 | 6.0 | 2 | 0.049 | 0.073 |
| MoCA.model = ATN_PD | 12 | 29,351 | -14,663 | 9.5 | 2 | 0.009 | 0.024 |
| SDMT.model = A | 8 | 28,729 | -14,356 |  |  |  |  |
| SDMT.model = AT_PD | 10 | 28,722 | -14,351 | 10.7 | 2 | 0.005 | 0.017 |
| SDMT.model = ATN_PD | 12 | 28,711 | -14,343 | 15.5 | 2 | <0.001 | 0.004 |
| Semantic Fluency.model = A | 8 | 28,573 | -14,278 |  |  |  |  |
| Semantic Fluency.model = AT_PD | 10 | 28,561 | -14,270 | 16.1 | 2 | <0.001 | 0.004 |
| Semantic Fluency.model = ATN_PD | 12 | 28,551 | -14,264 | 13.8 | 2 | 0.001 | 0.006 |
| UPDRS I cog.model = A | 8 | 29,291 | -14,637 |  |  |  |  |
| UPDRS I cog.model = AT_PD | 10 | 29,271 | -14,625 | 24.4 | 2 | <0.001 | <0.001 |
| UPDRS I cog.model = ATN_PD | 12 | 29,262 | -14,619 | 12.5 | 2 | 0.002 | 0.009 |
| Hoehn & Yahr.model = A | 8 | 22,880 | -11,432 |  |  |  |  |
| Hoehn & Yahr.model = AT_PD | 10 | 22,883 | -11,431 | 1.9 | 2 | 0.396 | 0.413 |
| Hoehn & Yahr.model = ATN_PD | 12 | 22,880 | -11,428 | 6.7 | 2 | 0.035 | 0.064 |
| PIGD.model = A | 8 | 29,842 | -14,913 |  |  |  |  |
| PIGD.model = AT_PD | 10 | 29,840 | -14,910 | 5.8 | 2 | 0.054 | 0.077 |
| PIGD.model = ATN_PD | 12 | 29,836 | -14,906 | 8.7 | 2 | 0.013 | 0.029 |
| SCOPA-AUT.model = A | 8 | 28,656 | -14,320 |  |  |  |  |
| SCOPA-AUT.model = AT_PD | 10 | 28,655 | -14,318 | 4.5 | 2 | 0.106 | 0.134 |
| SCOPA-AUT.model = ATN_PD | 12 | 28,653 | -14,315 | 6.1 | 2 | 0.048 | 0.073 |
| Tremor.model = A | 8 | 29,746 | -14,865 |  |  |  |  |
| Tremor.model = AT_PD | 10 | 29,748 | -14,864 | 2.3 | 2 | 0.318 | 0.347 |
| Tremor.model = ATN_PD | 12 | 29,749 | -14,863 | 3.1 | 2 | 0.217 | 0.248 |
| UPDRS III Motor.model = A | 8 | 23,056 | -11,520 |  |  |  |  |
| UPDRS III Motor.model = AT_PD | 10 | 23,055 | -11,517 | 4.9 | 2 | 0.088 | 0.117 |
| UPDRS III Motor.model = ATN_PD | 12 | 23,055 | -11,516 | 3.7 | 2 | 0.154 | 0.185 |

For traditional ATN (Supplemental Table 9), T status improved model fit for MoCA, HVLT, SDMT, Semantic Fluency, and MDS-UPDRS I Cog (all FDR-*p*<0.029). Results were not significant for JOLO (FDR-*p*=0.083), SCOPA-AUT (FDR-*p*=0.083), Letter Number Sequencing or any motor test (all FDR-*p*>0.16). Addition of N status using CSF t-tau did not improve any of the models (Letter Number Sequencing FDR-*p*=0.083; all other FDR-*p*>0.1).

## 4. Discussion

Here we tested ATN_PD_ – combining CSF Aβ_42_, p-tau_181_, and serum NfL – as a modified strategy to detect co-occurring AD in PD. Previous work shows that CSF Aβ_42_ predicts clinical outcomes in PD^17, 18, 35, 36^, while CSF p-tau_181_ and t-tau appear to have less utility in PD^18, 37^. This may be in part because CSF p-tau_181_ and t-tau levels are consistently low in PD^17^, which can make it difficult to determine clinically relevant levels of p-tau_181_. Here we tested if T status had added clinical utility to CSF Aβ_42_, despite low levels of CSF p-tau_181_ in PD. We also tested serum NfL as a marker of N, since NfL may be clinically useful in PD^8–11, 38^, although few have tested the contribution of NfL in combination with AD biomarkers in PD.

Our data show that ATN_PD_ classified 32 PD (9%) as AD positive (A+T+N±) at baseline. While small, this subset had strong associations with later cognitive decline; A+T+N± PD had more severe decline for all seven cognitive tests measured, with four significant after FDR-correction (MoCA, SDMT, Semantic Fluency, and MDS-UPDRS I Cog). These findings suggest an added predictive value of T status to A+ status in PD. While metrics of motor and autonomic functioning all significantly declined with time, only PIGD showed greater declines associated with A+T+N±. PIGD has been previously associated with CSF Aβ ^39, 40^. This specificity of A+T+N± with decline in cognitive function, but not motor or autonomic functioning, indicates that ATN_PD_ classifications are not simply capturing overall disease severity. A status alone was associated with some metrics of cognitive decline, still model comparisons showed that fitness significantly improved with ATN_PD_ and the addition of T status, and again with N status. Figure 5 highlights the divergent trajectories of A+T+N± (“AD”) from A+T-N-(“Amyloid”), further supporting that T and N statuses have added value for clinical prognosis (note also, A+T-N+, but with small sample size n=9). Importantly, we used a PD-specific cutpoint for p-tau_181_ (≥13 pg/mL)^19^, however results showed that p-tau_181_ also had prognostic utility when using the AD-derived cutpoint (≥24 pg/mL)^27^ (traditional ATN); thus our findings are not solely driven by the PD-specific cut point used to define ATN_PD_. In sum, CSF Aβ_42_, p-tau_181_, and serum NfL used in combination in ATN_PD_ may be highly informative for predicting clinical outcomes in PD.

Our examination of analyte trajectories confirms a biomarker profile in PD that is distinct from canonical AD, although the mechanism for this difference is unknown. Consistent with β-amyloid pathology (*i.e.*, A+ status), CSF Aβ_42_ was lower in PD than controls, and decreased over time. However, both CSF p-tau_181_ and t-tau were lower overall in PD than controls. This is in contrast to AD, where CSF p-tau_181_ and t-tau increase in association with tau pathology and neurodegeneration^41, 42^. Another distinction of these data is that CSF Aβ_42_, p-tau_181_ and t-tau were all positively correlated. Again, this pattern is in contrast to AD studies which instead show an inverse association between CSF Aβ_42_ and p-tau_181_ and t-tau (low Aβ_42_ and high p-tau_181_/t-tau)^43^. We tested how biomarker trajectories might differ by baseline CSF Aβ_42_, hypothesizing that CSF p-tau_181_ and t-tau may show greater increases in PD with low baseline CSF Aβ_42_ (A+). Overall CSF p-tau_181_ and t-tau were lower in PD with low CSF Aβ_42_, and longitudinal models showed that CSF p-tau_181_ and t-tau significantly decreased over time in PD with higher CSF Aβ_42_ levels (A-). It is possible that early PD is marked by initial declines in CSF tau, and future studies are needed to test if CSF tau rises in late-stage PD. The low tau levels observed here are in agreement with autopsy studies showing that tau pathology is less abundant in PD and other LBD than AD^44^. Further, CSF p-tau_181_ and t-tau may have an inverse association with α-synuclein pathology^15^. Still, the mechanism for low CSF levels in PD is unknown and our analyses showed no interaction between baseline CSF α-syn and AD biomarkers. Given this altered CSF profile in PD, we developed a modified ATN_PD_ strategy, which applied a lower, PD-established threshold for p-tau_181_^19^. We observed that CSF p-tau_181_ is informative for predicting cognitive decline in PD, and models demonstrated improved fit when combining CSF p-tau_181_ with Aβ_42_. Moreover, our results were robust and not cutpoint dependent: traditional ATN with a higher p-tau_181_ cutpoint also predicted cognitive decline, albeit for a smaller proportion of PD (n=7; 2%). This is corroborated by other studies that show improved predictive accuracy for cognitive decline in PD when combining CSF Aβ_42_ and p-tau ^45^. In sum, our results indicate that CSF p-tau_181_ has added utility when combined with CSF Aβ_42_.

In addition to threshold adjustments for CSF p-tau_181_, ATN_PD_ incorporated serum NfL as a marker of N. NfL is a structural component of myelinated axons, and concentrations of NfL increase in the blood and CSF following axonal injury^46^. The application of NfL in PD and in the ATN_PD_ framework has several advantages. First, unlike CSF t-tau, we show that serum NfL is not correlated with CSF Aβ_42_ or p-tau_181_, and is invariant to other indicators of AD pathology including APOE ε4; thus, serum NfL is an orthogonal marker of N with increased utility compared to t-tau. Our results showed improved model fit for 7 of 12 clinical outcomes with inclusion of N status using serum NfL under ATN_PD_. In contrast, traditional ATN using t-tau as a marker of N status did not improve model fit. CSF t-tau’s lack of added utility is in part because it is highly correlated with p-tau_181_, both in this PD cohort (rho=0.97; Supplementary Figure 1) and in AD samples^47^; thus t-tau is somewhat redundant with p-tau_181_. Second, there is substantial evidence that NfL is a more sensitive marker of N for non-AD neurodegenerative diseases than t-tau^48^, and strongly associates with disease progression, motor and cognitive severity in PD^38, 49, 50^. In this study, we also find that NfL levels significantly increased with time, and this increase was greater in PD than Controls. The utility of serum NfL in PD was further supported by improved model fit, not just for cognitive outcomes, but also for autonomic (SCOPA-AUT) and some motor outcomes (PIGD, Hoehn & Yahr), although only PIGD survived FDR-correction. Our data indicate that serum NfL does not associate with AD biomarkers in PD, and thus the predictive advantage of serum NfL in PD may possibly be due to a closer association with α-synuclein pathological processes, however this potential association needs to be confirmed.

There are several caveats to consider when interpreting our findings here. Foremost, while our results firmly show that ATN_PD_ is clinically relevant, we lack autopsy data to test external validity of ATN_PD_ to identify co-occurring AD compared to other biomarker strategies. This limitation highlights the need to test PD-specific biomarker cutpoints using autopsy data. Even so, detailed, systematic, longitudinal data in PD are rare, and because PPMI is an international multi-center study, it is more robust against selection and collection bias. Thus we find strong evidence that the combination of CSF Aβ_42_, p-tau_181_, and serum NfL are useful for predictive cognitive decline in PD. Second, there is minimal overall cognitive impairment in PPMI during this time interval of follow up. Likewise, only 32 PD met an ATN_PD_ definition of AD (A+T+N±) at baseline. While our findings indicate that these A+T+N± individuals had a divergent cognitive trajectory, it will be important to validate these findings in a larger sample of A+T+N± PD, beyond the 5 years in this study. Future studies should test the prognostic accuracy of ATN_PD_ categories to predict which PD individuals will develop overt dementia, or to distinguish fast *vs*. slow progressors. It will also be important to determine the optimal level of classification detail that best predicts clinical outcomes in PD in future work (*e.g.*, A+SNAP [A+T-N+] *vs* AD [A+T+N±]) since we were limited in numbers for some subgroups of ATN classification (A+SNAP n=9). Third, we did not test alternative strategies combining CSF Aβ_42_ and p-tau_181_, such as the p-tau_181_/Aβ_42_, which has been shown to contribute to predictions of cognitive decline in PD^45^. Fourth, we did not test application in other LBD, such as dementia with lewy bodies (DLB) and PD with dementia (PDD), and it will be important to test if ATN_PD_ broadly applies to α-synucleinopathies. Finally, we lacked an accurate marker of α-synuclein pathology to incorporate in ATN_PD_. Previous work shows poor prognosis of motor function in PD using available CSF α-syn^22^. Recent seeding assay technology has made possible accurate stratification using CSF α-synuclein, and future longitudinal studies in prodromal and early PD should test the trajectories of ATN_PD_ + α-synuclein to understand the temporal dynamics and interactions between biomarkers.

In sum, this paper shows that an ATN_PD_ strategy which combines AD CSF biomarkers with serum NfL can classify early PD into biological subgroups that show clinically meaningful differences within 5 years. This study also highlights the need to study AD co-pathology and biomarkers in a PD-specific context, including continued tracking of AD biomarkers in across disease stage in PD and the establishment of PD-specific cutpoints for AD biomarkers using autopsy-confirmed data.

## Acknowledgement

This work is supported by funding from the National Institute of Aging (P30-AG072979). KAQC is supported by the Alzheimer’s Association (AARF-D-619473, AARF-D-619473-RAPID). PPMI is sponsored by the Michael J. Fox Foundation for Parkinson’s Research (MJFF) and is co-funded by MJFF, Abbvie, Allergan, Avid Radiopharmaceuticals, Biogen, BioLegend, Bristol-Myers Squibb, Celgene, Denali, Eli Lilly & Co., F. Hoffman-La Roche, Ltd., GE Healthcare, Genentech, GlaxoSmithKline, Lundbeck, Merck, Meso-Scale, Piramal, Prevail Therapeutics, Pfizer, Roche, Sanofi Genzyme, Servier, Takeda, Teva, UCB, Berily, and Voyager Therapeutics. This work was supported by a grant from MJFF.

## Author Contributions

KC, DI, TT, ER, JP, AC, DI, and LS all contributed to the methodology and design of the study. KC, DI, CC, JK, TS, TF, AT, CT, KK, BM, DG, SH, DW, AS, KM, KP, and LS contributed to the acquisition and analysis of data. KC contributed to drafting the text or preparing the figures.

## Potential Conflicts of Interest

LMS received support in an investigator-initiated grant from Roche for all immunoassay reagents and instrumentation for the analyses in CSF samples of Aβ_42_, t-tau and ptau_181_. Authors report no conflicts of interest relevant to this study.

## Notes

### Author Declarations

The Parkinson's Progression Marker Initiative (PPMI) is a longitudinal, observational study, with data collection from approximately 50 international sites. Data were selected on September 7, 2021 from the PPMI database. All procedures were performed with prior approval from ethical standards committees at each participating institution and with informed consent from all study participants.

