## Supplemental Figures and Tables for "Evaluation of ATN_PD_ framework and biofluid markers to predict cognitive decline in early Parkinson’s disease"

### Supplemental Material

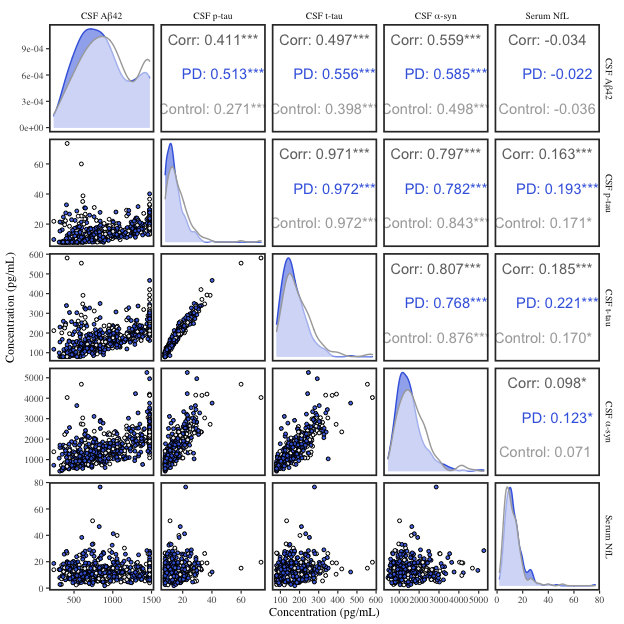

###### **Supplementary Figure 1.** **Pairwise analyte correlations at baseline**. Scatterplots and density distributions for each analyte are plotted for both PD (blue; n=364) and Controls (white; n=168). Spearman’s correlations were performed for the full group (Corr), PD, and Controls, with rho correlation coefficient reported. Asterisks represent nominal *p*-values from correlations (* *p*<0.05, ** *p*<0.01, *** *p*<0.001, with no asterisks indicating not significant (*p*>0.05).

| **CSF Aβ42** | **β** | | **95%CI** | | **Df** | | **p** |
| --- | --- | --- | --- | --- | --- | --- | --- |
| PD | -0.091 | | -0.163 -- -0.018 | | 554.0 | | 0.0153 |
| Visit Year | -0.007 | | -0.015 -- 0.001 | | 1,699.8 | | 0.0888 |
| Age at Baseline | -0.002 | | -0.005 -- 0.001 | | 486.7 | | 0.2133 |
| Gender = Female | 0.036 | | -0.033 -- 0.105 | | 483.9 | | 0.3065 |
| APOE ε4 | -0.280 | | -0.354 -- -0.206 | | 484.2 | | <0.0001 |
| PD:Visit Year | -0.010 | | -0.02 -- 0 | | 1,699.8 | | 0.0457 |
| **CSF p-tau** | **β** | | **95%CI** | | **Df** | | **p** |
| PD | -0.161 | | -0.229 -- -0.092 | | 511.1 | | <0.0001 |
| Visit Year | 0.003 | | -0.003 -- 0.008 | | 1,680.2 | | 0.2993 |
| Age at Baseline | 0.012 | | 0.009 -- 0.015 | | 480.9 | | <0.0001 |
| Gender = Female | 0.081 | | 0.015 -- 0.147 | | 479.5 | | 0.0172 |
| APOE ε4 | 0.041 | | -0.03 -- 0.112 | | 479.7 | | 0.2585 |
| PD:Visit Year | -0.004 | | -0.01 -- 0.003 | | 1,680.3 | | 0.2993 |
| **CSF t-tau** | **β** | | **95%CI** | | **Df** | | **p** |
| PD | -0.129 | | -0.193 -- -0.065 | | 533.7 | | 1e-04 |
| Visit Year | 0.005 | | -0.001 -- 0.012 | | 1,694.3 | | 0.0958 |
| Age at Baseline | 0.012 | | 0.009 -- 0.014 | | 483.1 | | <0.0001 |
| Gender = Female | 0.090 | | 0.029 -- 0.152 | | 481.0 | | 0.0042 |
| APOE ε4 | 0.026 | | -0.04 -- 0.092 | | 481.2 | | 0.4358 |
| PD:Visit Year | -0.005 | | -0.012 -- 0.003 | | 1,694.4 | | 0.2392 |
| **Serum NfL** | **β** | **95%CI** | | **Df** | | **p** | |
| PD | 0.060 | -0.007 -- 0.127 | | 657.9 | | 0.0804 | |
| Visit Year | 0.036 | 0.025 -- 0.047 | | 1,568.0 | | <0.0001 | |
| Age at Baseline | 0.035 | 0.032 -- 0.037 | | 472.2 | | <0.0001 | |
| Gender = Female | 0.113 | 0.053 -- 0.174 | | 473.9 | | 3e-04 | |
| APOE ε4 | 0.024 | -0.041 -- 0.09 | | 477.4 | | 0.462 | |
| PD:Visit Year | 0.035 | 0.022 -- 0.048 | | 1,570.0 | | <0.0001 | |

###### **Supplemental Table 1: Biofluid trajectories over time in Controls vs. PD.** Linear mixed effects models tested for an interaction between group (PD, Control) and change in biofluid over visit year (0-5). β-estimates, 95% confidence intervals for β-estimates (95%CI), and nominal *p*-values are reported.

| **CSF p-tau** | **β** | **95%CI** | | **Df** | | **p** |
| --- | --- | --- | --- | --- | --- | --- |
| Baseline CSF Aβ42 | 0.506 | 0.426 -- 0.587 | | 351.0 | | <0.0001 |
| Visit Year | 0.172 | 0.111 -- 0.232 | | 1,143.2 | | <0.0001 |
| Age at Baseline | 0.012 | 0.009 -- 0.016 | | 325.8 | | <0.0001 |
| Gender = Female | 0.042 | -0.023 -- 0.107 | | 325.8 | | 0.2057 |
| APOE ε4 | 0.121 | 0.049 -- 0.194 | | 325.4 | | 0.0012 |
| Baseline Aβ42:Visit Year | -0.026 | -0.035 -- -0.017 | | 1,143.1 | | <0.0001 |
| **CSF t-tau** | **β** | **95%CI** | | **Df** | | **p** |
| Baseline CSF Aβ42 | 0.503 | 0.428 -- 0.578 | | 375.9 | | <0.0001 |
| Visit Year | 0.165 | 0.092 -- 0.239 | | 1,158.5 | | <0.0001 |
| Age at Baseline | 0.012 | 0.009 -- 0.015 | | 328.2 | | <0.0001 |
| Gender = Female | 0.053 | -0.006 -- 0.113 | | 328.2 | | 0.081 |
| APOE ε4 | 0.111 | 0.044 -- 0.177 | | 327.4 | | 0.0012 |
| Baseline Aβ42:Visit Year | -0.025 | -0.035 -- -0.014 | | 1,158.5 | | <0.0001 |
| **Serum NfL** | **β** | **95%CI** | **Df** | | **p** | |
| Baseline CSF Aβ42 | -0.006 | -0.106 -- 0.094 | 431.0 | | 0.9112 | |
| Visit Year | 0.093 | -0.034 -- 0.221 | 1,067.5 | | 0.1522 | |
| Age at Baseline | 0.036 | 0.032 -- 0.04 | 320.5 | | <0.0001 | |
| Gender = Female | 0.125 | 0.049 -- 0.202 | 322.9 | | 0.0015 | |
| APOE ε4 | 0.045 | -0.041 -- 0.13 | 324.6 | | 0.3102 | |
| Baseline Aβ42:Visit Year | -0.003 | -0.022 -- 0.016 | 1,067.3 | | 0.7308 | |

###### **Supplemental Table 2: Biofluid trajectories by baseline CSF Aβ_42_ in PD.** In PD, linear mixed effects models tested for an interaction between CSF Aβ_42_ and longitudinal changes in CSF p-tau_181_, t-tau, and serum NfL. β-estimates, 95% confidence intervals for β-estimates (95%CI), and nominal *p*-values are reported.

| **HVLT** | **β** | | | **95%CI** | | | | **Df** | | | | | **p** | | |
| --- | --- | --- | --- | --- | --- | --- | --- | --- | --- | --- | --- | --- | --- | --- | --- |
| ATN_PD = AD (A+T+N±) | -117.252 | | | -280.52 -- 45.995 | | | | 754.1 | | | | | 0.1608 | | |
| Visit Year | 48.404 | | | 36.585 -- 60.263 | | | | 1,639.7 | | | | | <0.0001 | | |
| Education (years) | 24.927 | | | 12.17 -- 37.684 | | | | 353.7 | | | | | 2e-04 | | |
| Gender = Female | 199.051 | | | 120.776 -- 277.325 | | | | 352.0 | | | | | <0.0001 | | |
| AD(A+T+N±):Visit Year | -57.833 | | | -100.475 -- -15.217 | | | | 1,668.5 | | | | | 0.0079 | | |
| **JOLO** | **β** | | | **95%CI** | | | | | **Df** | | | | | **p** | |
| ATN_PD = AD (A+T+N±) | 6.572 | | | -164.987 -- 178.105 | | | | | 605.3 | | | | | 0.9403 | |
| Visit Year | -2.597 | | | -13.289 -- 8.15 | | | | | 1,623.9 | | | | | 0.6347 | |
| Education (years) | 37.804 | | | 23.534 -- 52.074 | | | | | 356.2 | | | | | <0.0001 | |
| Gender = Female | -229.004 | | | -316.638 -- -141.375 | | | | | 354.8 | | | | | <0.0001 | |
| AD(A+T+N±):Visit Year | -52.605 | | | -91.95 -- -13.266 | | | | | 1,652.7 | | | | | 0.0089 | |
| **Letter Number Sequencing** | | **β** | | | **95%CI** | | | | | **Df** | | | | | **p** |
| ATN_PD = AD (A+T+N±) | | -212.550 | | | -397.933 -- -27.173 | | | | | 483.7 | | | | | 0.0255 |
| Visit Year | | -20.233 | | | -29.065 -- -11.38 | | | | | 1,620.4 | | | | | <0.0001 |
| Education (years) | | 27.700 | | | 11.281 -- 44.119 | | | | | 362.5 | | | | | 0.0011 |
| Gender = Female | | 91.403 | | | -9.47 -- 192.269 | | | | | 361.0 | | | | | 0.0774 |
| AD(A+T+N±):Visit Year | | -39.048 | | | -70.89 -- -7.22 | | | | | 1,631.9 | | | | | 0.0163 |
| **MoCA** | **β** | | | **95%CI** | | | | **Df** | | | | | **p** | | |
| ATN_PD = AD (A+T+N±) | -122.719 | | | -302.205 -- 56.754 | | | | 534.8 | | | | | 0.182 | | |
| Visit Year | 3.404 | | | -6.352 -- 13.178 | | | | 1,625.1 | | | | | 0.4946 | | |
| Education (years) | 26.228 | | | 10.753 -- 41.704 | | | | 361.2 | | | | | 0.001 | | |
| Gender = Female | 220.002 | | | 124.942 -- 315.064 | | | | 359.8 | | | | | <0.0001 | | |
| AD(A+T+N±):Visit Year | -72.624 | | | -108.159 -- -37.121 | | | | 1,645.7 | | | | | 1e-04 | | |
| **SDMT** | **β** | | | **95%CI** | | | **Df** | | | | | **p** | | | |
| ATN_PD = AD (A+T+N±) | -173.981 | | | -359.947 -- 11.982 | | | 452.4 | | | | | 0.0682 | | | |
| Visit Year | -14.478 | | | -22.321 -- -6.612 | | | 1,615.3 | | | | | 3e-04 | | | |
| Education (years) | 33.976 | | | 17.224 -- 50.728 | | | 362.3 | | | | | 1e-04 | | | |
| Gender = Female | 225.979 | | | 123.037 -- 328.917 | | | 361.1 | | | | | <0.0001 | | | |
| AD(A+T+N±):Visit Year | -59.851 | | | -88.202 -- -31.532 | | | 1,625.1 | | | | | <0.0001 | | | |
| **Semantic Fluency** | **β** | | | **95%CI** | | | | **Df** | | | | | **p** | | |
| ATN_PD = AD (A+T+N±) | -100.313 | | | -281.809 -- 81.179 | | | | 445.7 | | | | | 0.2807 | | |
| Visit Year | -3.930 | | | -11.418 -- 3.575 | | | | 1,612.9 | | | | | 0.3043 | | |
| Education (years) | 41.425 | | | 25.033 -- 57.817 | | | | 360.5 | | | | | <0.0001 | | |
| Gender = Female | 356.181 | | | 255.45 -- 456.912 | | | | 359.4 | | | | | <0.0001 | | |
| AD(A+T+N±):Visit Year | -73.475 | | | -100.505 -- -46.495 | | | | 1,621.9 | | | | | <0.0001 | | |
| **MDS-UPDRS I Cog** | **β** | | **95%CI** | | | **Df** | | | | | **p** | | | | |
| ATN_PD = AD (A+T+N±) | 197.868 | | 52.956 -- 342.817 | | | 674.5 | | | | | 0.0078 | | | | |
| Visit Year | 37.751 | | 27.995 -- 47.488 | | | 1,647.0 | | | | | <0.0001 | | | | |
| Education (years) | -11.847 | | -23.57 -- -0.122 | | | 358.3 | | | | | 0.0491 | | | | |
| Gender = Female | -73.636 | | -145.615 -- -1.653 | | | 357.1 | | | | | 0.0463 | | | | |
| AD(A+T+N±):Visit Year | 65.070 | | 29.849 -- 100.283 | | | 1,667.8 | | | | | 3e-04 | | | | |

###### **Supplemental Table 3: Modified ATN_PD_ prediction of cognitive trajectories.** Linear mixed effect models tested for an interaction between ATN_PD_ status at baseline (AD [A+T+N±] *vs*. not [all other statuses]) and change in each clinical outcome. β-estimates, 95% confidence intervals for β-estimates (95%CI), and nominal *p*-values are reported.

| **HVLT** | **β** | | | | **95%CI** | | | | | **Df** | | | | | **p** | | | | |
| --- | --- | --- | --- | --- | --- | --- | --- | --- | --- | --- | --- | --- | --- | --- | --- | --- | --- | --- | --- |
| A Status = A+ | -40.057 | | | | -138.962 -- 58.827 | | | | | 733.5 | | | | | 0.4288 | | | | |
| Visit Year | 49.272 | | | | 35.521 -- 63.06 | | | | | 1,639.7 | | | | | <0.0001 | | | | |
| Education (years) | 21.882 | | | | 9.057 -- 34.707 | | | | | 353.7 | | | | | 9e-04 | | | | |
| Gender = Female | 190.703 | | | | 111.333 -- 270.071 | | | | | 352.7 | | | | | <0.0001 | | | | |
| A+:Visit Year | -16.148 | | | | -40.632 -- 8.353 | | | | | 1,645.2 | | | | | 0.1965 | | | | |
| **JOLO** | **β** | | | | | **95%CI** | | | | | | **Df** | | | | | **p** | | |
| A Status = A+ | -41.191 | | | | | -144.111 -- 61.709 | | | | | | 600.8 | | | | | 0.4344 | | |
| Visit Year | 0.998 | | | | | -11.419 -- 13.467 | | | | | | 1,623.8 | | | | | 0.8751 | | |
| Education (years) | 36.415 | | | | | 22.304 -- 50.524 | | | | | | 355.6 | | | | | <0.0001 | | |
| Gender = Female | -234.468 | | | | | -321.853 -- -147.088 | | | | | | 354.7 | | | | | <0.0001 | | |
| A+:Visit Year | -23.748 | | | | | -45.99 -- -1.49 | | | | | | 1,630.8 | | | | | 0.0366 | | |
| **Letter Number Sequencing** | | | | **β** | | | | | **95%CI** | | | | | **Df** | | | | | **p** |
| A Status = A+ | | | | -3.718 | | | | | -116.822 -- 109.38 | | | | | 477.4 | | | | | 0.9488 |
| Visit Year | | | | -17.295 | | | | | -27.56 -- -7.011 | | | | | 1,620.4 | | | | | 0.001 |
| Education (years) | | | | 24.045 | | | | | 7.512 -- 40.578 | | | | | 362.4 | | | | | 0.0047 |
| Gender = Female | | | | 82.819 | | | | | -19.589 -- 185.221 | | | | | 361.3 | | | | | 0.1149 |
| A+:Visit Year | | | | -18.507 | | | | | -36.784 -- -0.223 | | | | | 1,622.3 | | | | | 0.0474 |
| **MoCA** | **β** | | | | **95%CI** | | | | | **Df** | | | | | **p** | | | | |
| A Status = A+ | 39.390 | | | | -70.086 -- 148.857 | | | | | 523.9 | | | | | 0.4822 | | | | |
| Visit Year | 7.850 | | | | -3.499 -- 19.209 | | | | | 1,624.9 | | | | | 0.1756 | | | | |
| Education (years) | 22.725 | | | | 7.11 -- 38.34 | | | | | 361.3 | | | | | 0.0047 | | | | |
| Gender = Female | 212.981 | | | | 116.272 -- 309.693 | | | | | 360.3 | | | | | <0.0001 | | | | |
| A+:Visit Year | -31.127 | | | | -51.375 -- -10.852 | | | | | 1,628.9 | | | | | 0.0026 | | | | |
| **SDMT** | **β** | | | | **95%CI** | | | | | **Df** | | | | | **p** | | | | |
| A Status = A+ | -51.575 | | | | -165.003 -- 61.85 | | | | | 448.6 | | | | | 0.3747 | | | | |
| Visit Year | -14.390 | | | | -23.538 -- -5.222 | | | | | 1,615.5 | | | | | 0.0021 | | | | |
| Education (years) | 30.267 | | | | 13.424 -- 47.111 | | | | | 362.2 | | | | | 5e-04 | | | | |
| Gender = Female | 215.951 | | | | 111.604 -- 320.296 | | | | | 361.3 | | | | | 1e-04 | | | | |
| A+:Visit Year | -14.588 | | | | -30.888 -- 1.716 | | | | | 1,617.1 | | | | | 0.0797 | | | | |
| **Semantic Fluency** | | **β** | | | | | **95%CI** | | | | **Df** | | | | | **p** | | | |
| A Status = A+ | | -4.072 | | | | | -114.824 -- 106.678 | | | | 442.9 | | | | | 0.9428 | | | |
| Visit Year | | -4.790 | | | | | -13.549 -- 3.989 | | | | 1,613.5 | | | | | 0.2845 | | | |
| Education (years) | | 38.197 | | | | | 21.715 -- 54.679 | | | | 360.7 | | | | | <0.0001 | | | |
| Gender = Female | | 348.889 | | | | | 246.78 -- 450.997 | | | | 359.8 | | | | | <0.0001 | | | |
| A+:Visit Year | | -14.966 | | | | | -30.575 -- 0.63 | | | | 1,614.9 | | | | | 0.0603 | | | |
| **MDS-UPDRS I Cog** | | | **β** | | | | | **95%CI** | | | | | **Df** | | | | | **p** | |
| A Status = A+ | | | 27.926 | | | | | -61.177 -- 117.043 | | | | | 649.6 | | | | | 0.5404 | |
| Visit Year | | | 31.983 | | | | | 20.657 -- 43.292 | | | | | 1,645.7 | | | | | <0.0001 | |
| Education (years) | | | -7.509 | | | | | -19.492 -- 4.474 | | | | | 358.1 | | | | | 0.2215 | |
| Gender = Female | | | -61.733 | | | | | -135.937 -- 12.475 | | | | | 357.6 | | | | | 0.1049 | |
| A+:Visit Year | | | 33.407 | | | | | 13.207 -- 53.581 | | | | | 1,650.0 | | | | | 0.0012 | |

###### **Supplemental Table 4: A Status prediction of cognitive trajectories.** Linear mixed effect models tested for an interaction between A status at baseline (A+ *vs*. A-) and change in each clinical outcome. β-estimates, 95% confidence intervals for β-estimates (95%CI), and nominal *p*-values are reported.

| **HVLT** | **β** | | | **95%CI** | | | | **Df** | | | | | **p** | | |
| --- | --- | --- | --- | --- | --- | --- | --- | --- | --- | --- | --- | --- | --- | --- | --- |
| ATN = AD (A+T+N±) | -22.028 | | | -357.1 -- 312.997 | | | | 728.8 | | | | | 0.8978 | | |
| Visit Year | 46.834 | | | 35.392 -- 58.316 | | | | 1,642.9 | | | | | <0.0001 | | |
| Education (years) | 23.201 | | | 10.375 -- 36.027 | | | | 353.8 | | | | | 5e-04 | | |
| Gender = Female | 200.538 | | | 121.307 -- 279.767 | | | | 352.8 | | | | | <0.0001 | | |
| AD(A+T+N±):Visit Year | -142.488 | | | -226.713 -- -58.178 | | | | 1,609.1 | | | | | 9e-04 | | |
| **JOLO** | **β** | | | **95%CI** | | | | | **Df** | | | | | **p** | |
| ATN = AD (A+T+N±) | -214.425 | | | -565.458 -- 136.593 | | | | | 607.4 | | | | | 0.233 | |
| Visit Year | -5.263 | | | -15.645 -- 5.177 | | | | | 1,627.2 | | | | | 0.3217 | |
| Education (years) | 37.917 | | | 23.784 -- 52.05 | | | | | 355.7 | | | | | <0.0001 | |
| Gender = Female | -223.883 | | | -311.229 -- -136.543 | | | | | 354.6 | | | | | <0.0001 | |
| AD(A+T+N±):Visit Year | -69.949 | | | -151.252 -- 11.345 | | | | | 1,612.7 | | | | | 0.0919 | |
| **Letter Number Sequencing** | | **β** | | | **95%CI** | | | | | **Df** | | | | | **p** |
| ATN = AD (A+T+N±) | | 102.725 | | | -282.667 -- 488.106 | | | | | 475.3 | | | | | 0.6027 |
| Visit Year | | -21.658 | | | -30.22 -- -13.075 | | | | | 1,621.4 | | | | | <0.0001 |
| Education (years) | | 24.361 | | | 7.752 -- 40.972 | | | | | 362.4 | | | | | 0.0044 |
| Gender = Female | | 86.088 | | | -16.601 -- 188.772 | | | | | 361.4 | | | | | 0.1022 |
| AD(A+T+N±):Visit Year | | -79.293 | | | -142.049 -- -16.472 | | | | | 1,604.2 | | | | | 0.0134 |
| **MoCA** | **β** | | | **95%CI** | | | | **Df** | | | | | **p** | | |
| ATN = AD (A+T+N±) | -182.254 | | | -554.265 -- 189.76 | | | | 535.6 | | | | | 0.3388 | | |
| Visit Year | 0.077 | | | -9.406 -- 9.577 | | | | 1,627.6 | | | | | 0.9874 | | |
| Education (years) | 24.432 | | | 8.895 -- 39.969 | | | | 361.3 | | | | | 0.0023 | | |
| Gender = Female | 222.729 | | | 126.683 -- 318.777 | | | | 360.4 | | | | | <0.0001 | | |
| AD(A+T+N±):Visit Year | -107.032 | | | -177.103 -- -36.913 | | | | 1,603.8 | | | | | 0.0028 | | |
| **SDMT** | **β** | | | **95%CI** | | | | **Df** | | | | | **p** | | |
| ATN = AD (A+T+N±) | -381.515 | | | -764.364 -- 1.333 | | | | 448.8 | | | | | 0.0521 | | |
| Visit Year | -17.489 | | | -25.116 -- -9.84 | | | | 1,616.7 | | | | | <0.0001 | | |
| Education (years) | 32.393 | | | 15.654 -- 49.133 | | | | 362.2 | | | | | 2e-04 | | |
| Gender = Female | 230.164 | | | 126.658 -- 333.668 | | | | 361.3 | | | | | <0.0001 | | |
| AD(A+T+N±):Visit Year | -78.915 | | | -134.831 -- -23.007 | | | | 1,602.4 | | | | | 0.0057 | | |
| **Semantic Fluency** | **β** | | | **95%CI** | | | **Df** | | | | | **p** | | | |
| ATN = AD (A+T+N±) | -324.053 | | | -698.584 -- 50.473 | | | 442.9 | | | | | 0.0916 | | | |
| Visit Year | -8.342 | | | -15.651 -- -1.019 | | | 1,614.6 | | | | | 0.0256 | | | |
| Education (years) | 39.981 | | | 23.567 -- 56.396 | | | 360.7 | | | | | <0.0001 | | | |
| Gender = Female | 359.355 | | | 257.859 -- 460.851 | | | 359.9 | | | | | <0.0001 | | | |
| AD(A+T+N±):Visit Year | -61.980 | | | -115.507 -- -8.406 | | | 1,600.7 | | | | | 0.0234 | | | |
| **MDS-UPDRS I Cog** | **β** | | **95%CI** | | | **Df** | | | | | **p** | | | | |
| ATN = AD (A+T+N±) | 411.534 | | 112.643 -- 710.448 | | | 661.8 | | | | | 0.0073 | | | | |
| Visit Year | 40.888 | | 31.408 -- 50.343 | | | 1,649.1 | | | | | <0.0001 | | | | |
| Education (years) | -9.875 | | -21.654 -- 1.904 | | | 357.8 | | | | | 0.1022 | | | | |
| Gender = Female | -77.372 | | -150.171 -- -4.57 | | | 357.3 | | | | | 0.0385 | | | | |
| AD(A+T+N±):Visit Year | 89.058 | | 17.636 -- 160.43 | | | 1,636.8 | | | | | 0.0146 | | | | |

###### **Supplemental Table 5: Traditional ATN prediction of cognitive trajectories.** Linear mixed effect models tested for an interaction between ATN status at baseline (AD [A+T+N±] *vs*. not [all other statuses]) and change in each clinical outcome. β-estimates, 95% confidence intervals for β-estimates (95%CI), and nominal *p*-values are reported.

| **Hoehn & Yahr** | **β** | **95%CI** | | | **Df** | | | | **p** | | | |
| --- | --- | --- | --- | --- | --- | --- | --- | --- | --- | --- | --- | --- |
| ATN_PD = AD (A+T+N±) | 61.399 | -54.404 -- 177.214 | | | 658.5 | | | | 0.3005 | | | |
| Visit Year | 65.811 | 57.391 -- 74.156 | | | 1,335.9 | | | | <0.0001 | | | |
| Education (years) | 13.823 | 4.456 -- 23.189 | | | 345.5 | | | | 0.0042 | | | |
| Gender = Female | 11.324 | -46.614 -- 69.27 | | | 352.2 | | | | 0.7028 | | | |
| AD(A+T+N±):Visit Year | -4.516 | -36.114 -- 26.957 | | | 1,349.9 | | | | 0.779 | | | |
| **PIGD** | **β** | **95%CI** | | | | **Df** | | | | **p** | | |
| ATN_PD = AD (A+T+N±) | 21.215 | -150.107 -- 192.527 | | | | 634.8 | | | | 0.8088 | | |
| Visit Year | 78.786 | 67.779 -- 89.759 | | | | 1,646.8 | | | | <0.0001 | | |
| Education (years) | 0.733 | -13.375 -- 14.839 | | | | 360.1 | | | | 0.9192 | | |
| Gender = Female | 11.091 | -75.511 -- 97.701 | | | | 358.6 | | | | 0.8025 | | |
| AD(A+T+N±):Visit Year | 68.581 | 29.001 -- 108.093 | | | | 1,662.7 | | | | 7e-04 | | |
| **SCOPA-AUT** | **β** | | | **95%CI** | | | **Df** | | | | **p** | |
| ATN_PD = AD (A+T+N±) | 286.998 | | | 100.129 -- 473.862 | | | 463.4 | | | | 0.0028 | |
| Visit Year | 64.613 | | | 56.487 -- 72.73 | | | 1,606.2 | | | | <0.0001 | |
| Education (years) | -16.332 | | | -33.052 -- 0.387 | | | 361.7 | | | | 0.0571 | |
| Gender = Female | -20.106 | | | -122.978 -- 82.762 | | | 359.7 | | | | 0.7027 | |
| AD(A+T+N±):Visit Year | 27.335 | | | -1.816 -- 56.458 | | | 1,610.0 | | | | 0.0662 | |
| **Tremor** | **β** | | | **95%CI** | | **Df** | | | | **p** | | |
| ATN_PD = AD (A+T+N±) | 135.220 | | | -47.107 -- 317.555 | | 561.7 | | | | 0.1478 | | |
| Visit Year | 57.674 | | | 47.235 -- 68.107 | | 1,641.5 | | | | <0.0001 | | |
| Education (years) | 7.278 | | | -8.257 -- 22.815 | | 362.7 | | | | 0.3605 | | |
| Gender = Female | -56.384 | | | -151.801 -- 39.03 | | 361.2 | | | | 0.2489 | | |
| AD(A+T+N±):Visit Year | 1.101 | | | -36.489 -- 38.683 | | 1,654.6 | | | | 0.9542 | | |
| **MDS-UPDRS III Motor** | **β** | | **95%CI** | | | | | **Df** | | | | **p** |
| ATN_PD = AD (A+T+N±) | 46.126 | | -101.252 -- 193.524 | | | | | 520.4 | | | | 0.5411 |
| Visit Year | 89.009 | | 80.424 -- 97.552 | | | | | 1,296.4 | | | | <0.0001 |
| Education (years) | 9.433 | | -3.349 -- 22.214 | | | | | 357.0 | | | | 0.1501 |
| Gender = Female | -73.186 | | -152.074 -- 5.705 | | | | | 361.7 | | | | 0.0707 |
| AD(A+T+N±):Visit Year | 13.307 | | -18.946 -- 45.507 | | | | | 1,307.3 | | | | 0.4186 |

###### **Supplemental Table 6: Modified ATN_PD_ prediction of motor and autonomic trajectories.** Linear mixed effect models tested for an interaction between ATN_PD_ status at baseline (AD [A+T+N±] *vs*. not [all other statuses]) and change in each clinical outcome. β-estimates, 95% confidence intervals for β-estimates (95%CI), and nominal *p*-values are reported.

| **Hoehn & Yahr** | **β** | | **95%CI** | | | **Df** | | | **p** | | |
| --- | --- | --- | --- | --- | --- | --- | --- | --- | --- | --- | --- |
| A Status = A+ | 49.839 | | -19.044 -- 118.748 | | | 634.7 | | | 0.1579 | | |
| Visit Year | 66.191 | | 56.607 -- 75.693 | | | 1,325.8 | | | <0.0001 | | |
| Education (years) | 14.417 | | 5.146 -- 23.686 | | | 344.3 | | | 0.0026 | | |
| Gender = Female | 14.172 | | -43.685 -- 72.034 | | | 352.2 | | | 0.6325 | | |
| A+:Visit Year | -2.249 | | -20.17 -- 15.68 | | | 1,351.5 | | | 0.8058 | | |
| **PIGD** | **β** | | | **95%CI** | | | **Df** | | | **p** | |
| A Status = A+ | -61.390 | | | -164.814 -- 42.032 | | | 627.4 | | | 0.2464 | |
| Visit Year | 74.776 | | | 61.973 -- 87.53 | | | 1,646.4 | | | <0.0001 | |
| Education (years) | 3.008 | | | -11.045 -- 17.058 | | | 359.2 | | | 0.6759 | |
| Gender = Female | 15.430 | | | -71.565 -- 102.435 | | | 358.4 | | | 0.7291 | |
| A+:Visit Year | 29.147 | | | 6.378 -- 51.942 | | | 1,650.4 | | | 0.0123 | |
| **SCOPA-AUT** | **β** | | | **95%CI** | | | **Df** | | | **p** | |
| A Status = A+ | 63.846 | | | -50.095 -- 177.786 | | | 455.0 | | | 0.2741 | |
| Visit Year | 61.981 | | | 52.562 -- 71.391 | | | 1,604.4 | | | <0.0001 | |
| Education (years) | -12.064 | | | -28.895 -- 4.767 | | | 360.3 | | | 0.1621 | |
| Gender = Female | -8.662 | | | -113.092 -- 95.765 | | | 359.0 | | | 0.8713 | |
| A+:Visit Year | 14.880 | | | -1.95 -- 31.696 | | | 1,606.8 | | | 0.0832 | |
| **Tremor** | **β** | | | **95%CI** | | | **Df** | | | **p** | |
| A Status = A+ | -21.828 | | | -131.824 -- 88.171 | | | 555.3 | | | 0.6983 | |
| Visit Year | 52.132 | | | 40.031 -- 64.226 | | | 1,641.4 | | | <0.0001 | |
| Education (years) | 8.996 | | | -6.467 -- 24.458 | | | 362.1 | | | 0.2563 | |
| Gender = Female | -52.510 | | | -148.275 -- 43.251 | | | 361.2 | | | 0.2846 | |
| A+:Visit Year | 17.638 | | | -3.943 -- 39.214 | | | 1,644.5 | | | 0.1094 | |
| **MDS-UPDRS III Motor** | | **β** | | | **95%CI** | | | **Df** | | | **p** |
| A Status = A+ | | 5.129 | | | -83.09 -- 93.36 | | | 506.5 | | | 0.9096 |
| Visit Year | | 86.876 | | | 77.111 -- 96.584 | | | 1,290.9 | | | <0.0001 |
| Education (years) | | 10.298 | | | -2.384 -- 22.978 | | | 356.8 | | | 0.1134 |
| Gender = Female | | -70.895 | | | -149.834 -- 8.046 | | | 362.2 | | | 0.0801 |
| A+:Visit Year | | 10.938 | | | -7.417 -- 29.325 | | | 1,305.6 | | | 0.2435 |

###### **Supplemental Table 7: A Status prediction of motor and autonomic trajectories.** Linear mixed effect models tested for an interaction between A status at baseline (A+ *vs*. A-) and change in each clinical outcome. β-estimates, 95% confidence intervals for β-estimates (95%CI), and nominal nominal *p*-values are reported.

| **Hoehn & Yahr** | **β** | | **95%CI** | | **Df** | | | **p** | | |
| --- | --- | --- | --- | --- | --- | --- | --- | --- | --- | --- |
| ATN = AD (A+T+N±) | 160.606 | | -79.451 -- 400.697 | | 684.1 | | | 0.1915 | | |
| Visit Year | 65.648 | | 57.475 -- 73.742 | | 1,336.7 | | | <0.0001 | | |
| Education (years) | 13.990 | | 4.682 -- 23.298 | | 344.7 | | | 0.0035 | | |
| Gender = Female | 9.905 | | -48.037 -- 67.856 | | 351.6 | | | 0.7385 | | |
| AD(A+T+N±):Visit Year | -11.958 | | -87.126 -- 63.058 | | 1,346.9 | | | 0.755 | | |
| **PIGD** | **β** | | | **95%CI** | | | **Df** | | | **p** |
| ATN = AD (A+T+N±) | -133.958 | | | -485.018 -- 217.111 | | | 624.3 | | | 0.4561 |
| Visit Year | 81.040 | | | 70.373 -- 91.667 | | | 1,648.5 | | | <0.0001 |
| Education (years) | 2.101 | | | -11.968 -- 16.169 | | | 359.1 | | | 0.7705 |
| Gender = Female | 10.824 | | | -76.114 -- 97.772 | | | 358.3 | | | 0.8079 |
| AD(A+T+N±):Visit Year | 157.949 | | | 79.433 -- 236.446 | | | 1,621.3 | | | 1e-04 |
| **SCOPA-AUT** | **β** | | **95%CI** | | **Df** | | | **p** | | |
| ATN = AD (A+T+N±) | 484.299 | | 99.929 -- 868.667 | | 451.4 | | | 0.0142 | | |
| Visit Year | 66.257 | | 58.367 -- 74.134 | | 1,605.9 | | | <0.0001 | | |
| Education (years) | -14.075 | | -30.838 -- 2.688 | | 360.1 | | | 0.1017 | | |
| Gender = Female | -22.465 | | -126.286 -- 81.353 | | 358.8 | | | 0.6726 | | |
| AD(A+T+N±):Visit Year | 19.667 | | -37.929 -- 77.278 | | 1,591.5 | | | 0.5035 | | |
| **Tremor** | **β** | **95%CI** | | | | **Df** | | | **p** | |
| ATN = AD (A+T+N±) | -42.764 | -417.258 -- 331.742 | | | | 553.4 | | | 0.8235 | |
| Visit Year | 57.294 | 47.171 -- 67.408 | | | | 1,643.1 | | | <0.0001 | |
| Education (years) | 8.956 | -6.563 -- 24.474 | | | | 362.0 | | | 0.2601 | |
| Gender = Female | -53.296 | -149.226 -- 42.632 | | | | 361.2 | | | 0.2783 | |
| AD(A+T+N±):Visit Year | 20.433 | -54.144 -- 94.949 | | | | 1,620.2 | | | 0.5912 | |
| **MDS-UPDRS III Motor** | **β** | **95%CI** | | | | **Df** | | | **p** | |
| ATN = AD (A+T+N±) | -41.052 | -345.895 -- 263.798 | | | | 531.6 | | | 0.7925 | |
| Visit Year | 89.349 | 81.02 -- 97.631 | | | | 1,296.8 | | | <0.0001 | |
| Education (years) | 10.150 | -2.583 -- 22.881 | | | | 356.6 | | | 0.1201 | |
| Gender = Female | -72.329 | -151.408 -- 6.754 | | | | 361.5 | | | 0.0747 | |
| AD(A+T+N±):Visit Year | 48.102 | -28.588 -- 124.791 | | | | 1,303.3 | | | 0.2193 | |

###### **Supplemental Table 8: Traditional ATN prediction of motor and autonomic trajectories.** Linear mixed effect models tested for an interaction between ATN status at baseline (AD [A+T+N±] *vs*. not [all other statuses]) and change in each clinical outcome. β-estimates, 95% confidence intervals for β-estimates (95%CI), and nominal *p*-values are reported.

| Test | npar | AIC | logLik | χ2 | Df | p | FDR-p |
| --- | --- | --- | --- | --- | --- | --- | --- |
| MoCA.model = A | 8 | 29,358 | -14,671 |  |  |  |  |
| MoCA.model = AT | 10 | 29,346 | -14,663 | 16.7 | 2 | <0.001 | 0.003 |
| MoCA.model = ATN | 12 | 29,347 | -14,661 | 3.0 | 2 | 0.223 | 0.315 |
| HVLT.model = A | 8 | 29,794 | -14,889 |  |  |  |  |
| HVLT.model = AT | 10 | 29,785 | -14,882 | 13.4 | 2 | 0.001 | 0.009 |
| HVLT.model = ATN | 12 | 29,786 | -14,881 | 2.5 | 2 | 0.283 | 0.377 |
| SDMT.model = A | 8 | 28,729 | -14,356 |  |  |  |  |
| SDMT.model = AT | 10 | 28,723 | -14,351 | 10.2 | 2 | 0.006 | 0.029 |
| SDMT.model = ATN | 12 | 28,720 | -14,348 | 6.5 | 2 | 0.038 | 0.102 |
| Semantic Fluency.model = A | 8 | 28,573 | -14,278 |  |  |  |  |
| Semantic Fluency.model = AT | 10 | 28,564 | -14,272 | 13.0 | 2 | 0.002 | 0.009 |
| Semantic Fluency.model = ATN | 12 | 28,563 | -14,269 | 5.3 | 2 | 0.072 | 0.157 |
| Letter Number.model = A | 8 | 29,085 | -14,534 |  |  |  |  |
| Letter Number.model = AT | 10 | 29,084 | -14,532 | 4.5 | 2 | 0.104 | 0.192 |
| Letter Number.model = ATN | 12 | 29,081 | -14,529 | 7.2 | 2 | 0.028 | 0.083 |
| UPDRS I cog.model = A | 8 | 29,291 | -14,637 |  |  |  |  |
| UPDRS I cog.model = AT | 10 | 29,272 | -14,626 | 23.2 | 2 | <0.001 | <0.001 |
| UPDRS I cog.model = ATN | 12 | 29,272 | -14,624 | 3.8 | 2 | 0.150 | 0.252 |
| JOLO.model = A | 8 | 29,473 | -14,728 |  |  |  |  |
| JOLO.model = AT | 10 | 29,469 | -14,725 | 7.4 | 2 | 0.025 | 0.083 |
| JOLO.model = ATN | 12 | 29,470 | -14,723 | 3.5 | 2 | 0.175 | 0.262 |
| SCOPA-AUT.model = A | 8 | 28,656 | -14,320 |  |  |  |  |
| SCOPA-AUT.model = AT | 10 | 28,653 | -14,316 | 7.3 | 2 | 0.026 | 0.083 |
| SCOPA-AUT.model = ATN | 12 | 28,654 | -14,315 | 2.3 | 2 | 0.310 | 0.392 |
| UPDRS III Motor.model = A | 8 | 23,056 | -11,520 |  |  |  |  |
| UPDRS III Motor.model = AT | 10 | 23,054 | -11,517 | 5.3 | 2 | 0.069 | 0.157 |
| UPDRS III Motor.model = ATN | 12 | 23,058 | -11,517 | 1.0 | 2 | 0.616 | 0.665 |
| PIGD.model = A | 8 | 29,842 | -14,913 |  |  |  |  |
| PIGD.model = AT | 10 | 29,842 | -14,911 | 4.7 | 2 | 0.093 | 0.186 |
| PIGD.model = ATN | 12 | 29,845 | -14,910 | 0.8 | 2 | 0.665 | 0.665 |
| Hoehn & Yahr.model = A | 8 | 22,880 | -11,432 |  |  |  |  |
| Hoehn & Yahr.model = AT | 10 | 22,881 | -11,430 | 3.7 | 2 | 0.157 | 0.252 |
| Hoehn & Yahr.model = ATN | 12 | 22,883 | -11,430 | 1.5 | 2 | 0.474 | 0.542 |
| Tremor.model = A | 8 | 29,746 | -14,865 |  |  |  |  |
| Tremor.model = AT | 10 | 29,749 | -14,864 | 1.6 | 2 | 0.449 | 0.539 |
| Tremor.model = ATN | 12 | 29,752 | -14,864 | 0.9 | 2 | 0.645 | 0.665 |

###### **Supplemental Table 9: Comparison of nested models (A *vs*. AT *vs.* ATN) for prediction of clinical outcomes.** Summary of ANOVAs comparing model fitness for traditional ATN. Columns report number of parameters per model (npar), AIC, log liklihood (logLik), chi-square (χ^2^), degrees of freedom (Df), *p*-values, and FDR-adjusted *p*-values.
